# Integrated microbiome and metabolome analysis reveals the interaction between intestinal flora and serum metabolites as potential biomarkers in hepatocellular carcinoma patients

**DOI:** 10.1101/2022.12.24.22283879

**Authors:** Xiaoyue Li, Xinyu Gu, Liqiu Li, Liangliang Xiang, Zhaodi Jiang, Yufeng Zhang, Shujie Li, Changhua Yi, Junwei Li, Yongxiang Yi

## Abstract

Globally, liver cancer poses a serious threat to human health and quality of life. Despite numerous studies on the microbial composition of the gut in hepatocellular carcinoma (HCC), little is known about the interactions of the gut microbiota and metabolites and their role in HCC. We examined the composition of the gut microbiota and serum metabolic profiles in 68 patients with HCC, 33 patients with liver cirrhosis (LC), and 34 healthy individuals (NC) using a combination of metagenome sequencing and liquid chromatography‒mass spectrometry (LC‒MS). The composition of the serum metabolites and the structure of the intestinal microbiota were found to be significantly altered in HCC patients compared to non-HCC patients. We used LEfSe and metabolic pathway enrichment analysis to identify two key species (*Odoribacter splanchnicus* and *Ruminococcus bicirculans*) and five key metabolites (ouabain, taurochenodeoxycholic acid, glycochenodeoxycholate, theophylline, and xanthine) associated with HCC, which we then combined to create panels for HCC diagnosis. We discovered that the diagnostic performance of the metabolome was superior to that of the microbiome, and a panel comprised of key species and key metabolites outperformed AFP in terms of diagnostic value. Spearman’s rank correlation test was used to determine the relationship between the intestinal flora and serum metabolites and their impact on hepatocarcinogenesis and progression. A random forest model was used to assess the diagnostic performance of the different histologies alone and in combination. The panel we developed was very close to the AUC values of the best omics. In summary, this study describes the characteristics of HCC patients’ intestinal flora and serum metabolism, demonstrates that HCC is caused by the interaction of intestinal flora and serum metabolites, and suggests that two key species and five key metabolites may be potential markers for the diagnosis of HCC.

---

Primary liver cancer is the fourth leading cause of cancer-related deaths worldwide, with hepatocellular carcinoma (HCC) accounting for approximately 80% of all cases (1). China represents approximately half of all new cases and deaths related to HCC worldwide (2). Although surgery can help patients with liver cancer, the 5-year survival rate is only 50%–70% (3). Furthermore, most HCC patients are diagnosed in advanced stages with a poor prognosis due to a lack of specific symptoms in the early stages and no known early diagnostic markers (4). To screen for and diagnose HCC, imaging methods (e.g., CT and B-ultrasound) and serum biomarkers (e.g., alpha-fetoprotein [AFP]) are commonly used. However, imaging alone is insufficient for distinguishing small HCCs from hepatic sclerosing nodules. In addition, AFP has a sensitivity of 65% for the clinical diagnosis of HCC and a sensitivity of less than 40% for preclinical prediction (5). As a result, new biomarkers and effective drug targets are urgently needed to improve the prognosis of HCC patients.

Gut microbes and circulating metabolites have received much attention as biomarkers for human diseases such as cancer in recent years due to the development and application of sequencing technologies and LC‒MS-based metabolomics. Intestinal microbes are recognized as novel virtual metabolic organs, and the gut microbiota has been demonstrated to play a significant role in the development of numerous diseases. By altering the permeability of the intestinal mucosa in a way that disrupts immune or metabolic homeostasis, gut microbiota imbalances can contribute to the development of autoimmune diseases or cancer (6, 7, 8). The close relationship between the liver and the intestine is referred to as the “gut-liver axis” (9). An intact gut-liver axis is dependent on a healthy intestinal microbiota and normal liver function. In addition, the gut microbiota has been identified as an important player in chronic inflammatory liver disease, liver cirrhosis, alcoholic liver disease, and nonalcoholic fatty liver disease (10, 11, 12). Several studies have shown that the gut microbiota can be used as a noninvasive diagnostic tool for certain diseases and cancers, such as type 2 diabetes (T2D), colorectal cancer (CRC), and pancreatic cancer (PC) (13, 14, 15). Although the importance of microbes in HCC has been reported in several studies, the profile of the gut microbial community and its functional contribution to HCC has yet to be thoroughly studied and systematically characterized (16, 17).

Using microbial metabolites, the gut microbiota has been linked to diseases such as cancer (18). Food and nutrients are transformed by gut microbes into a metabolite environment, which controls the equilibrium of the metabolites (19). By providing metabolic flux to promote anabolism, acting as competitive enzyme inhibitors, or modifying signaling proteins, among other mechanisms, these metabolites can exert genotoxic or tumor-suppressive effects (20).

The liver is one of the most active metabolizing organs in our bodies, and it plays an important role in regulating various metabolic processes (21). The liver receives metabolites produced by bacteria in the gut via the portal vein and transports them directly to the liver to perform regulatory functions. Because of the natural connection between intestinal microbes and the liver, the liver is the first organ to receive intestinal metabolites and it plays an essential role in the interaction between extraneous materials and the systemic environment. Metabolomics is a very promising method for identifying metabolites that can shed light on the etiology, treatment, and early diagnosis of disease (22).

The process of tumorigenesis is accompanied by an overall shift in metabolic status, which has an effect not only on the tissue of the tumor but also on the microenvironment surrounding it (23, 24). Furthermore, metabolic changes can be observed more directly in the tumor cell state than genomic and proteomic changes and are thus expected to become useful tumorigenesis biomarkers (25). In the past few years, much research has been done on the metabolites in the blood that are linked to liver cancer. This research has shown that metabolites play a major role in the development of HCC (26, 27).

With increasing research on the gut microbiome and metabolome as biomarkers in HCC, we have a deeper understanding of possible diagnostic methods for HCC. However, little is known about the interactions between gut microbes and metabolites and how they influence the development of liver cancer. In this study, we performed metagenome sequencing of stool and metabolomic analysis of serum from three cohorts (HCC, LC, and NC) to discover changes in gut microbes and serum metabolites. Simultaneously, matched serum and stool samples were analyzed for metabolites and microbes, and it was discovered that gut microbes were closely associated with serum metabolites. Based on these findings, we compared the diagnostic performance of key gut microbes, key serum metabolites, and key gut microbial metabolites. Meanwhile, we examined the molecular pathway mechanisms to learn more about hepatocarcinogenesis.

## MATERIALS AND METHODS

### Participant information

This study included 68 newly diagnosed hepatocellular carcinoma patients, 33 patients with liver cirrhosis from the Hepatobiliary and Pancreatic Treatment Center of the Second Hospital of Nanjing, Jiangsu Province, and 34 healthy subjects from the Health Management Center. Stool and serum samples were collected in accordance with the protocol approved by the ethics committee of the Second Hospital of Nanjing, and all participants provided written informed consent. The study subjects’ demographic and clinicopathological data, CT scans, and dietary habits were obtained from hospital electronic medical records and questionnaires (online Supplementary Table S1). International guidelines say that HCC or cirrhosis can be diagnosed by looking at integrated pathology, imaging, laboratory tests, clinical symptoms, and medical history.

The HCC patients were screened and confirmed. The following were the exclusion criteria: 1) patients with other diseases, such as tumors in other locations, gastrointestinal diseases, hypertension, diabetes, and metabolic diseases; 2) patients who had previously received anticancer treatment; 3) patients who had recently received antibiotics or probiotics; and 4) patients who lacked critical clinical information. Age, sex ratio, and BMI were used to match the patients with the controls. Individuals who had hypertension, diabetes, obesity, metabolic syndrome, irritable bowel syndrome (IBD), or had received antibiotics and/or probiotics treatment within the previous 8 weeks were also excluded.

### Sample collection

Fresh fecal samples were collected from each participant and evaluated for fecal morphology and color. The samples were divided into three 300 mg portions and immediately snap-frozen in liquid nitrogen. The entire process of moving and manipulating the samples on ice took less than 20 minutes. After snap-freezing with liquid nitrogen, the samples were stored at −80 °C until extraction for testing. A professional nurse collected venous blood in strict accordance with standard asepsis procedures. The serum was centrifuged and stored at −80 °C until testing.

### Microbial DNA Extraction, Metagenome Sequencing and Data Processing

Total DNA was extracted from the stool samples using the QIAamp 96 Power Fecal QIAcube HT kit (Qiagen, Germany), and the DNA was further purified using the MGI Easy DNA Magnetic Beads Purification Kit (MGI, China) according to the manufacturer’s instructions. To measure how much purified DNA there was, a Qubit dsDNA BR Assay Kit (Invitrogen, USA) was used.

The library was built using DNBSEQ (online Supplementary Figure S1) and the original sequencing data (Raw Data) were filtered using the short oligonucleotide alignment program SOAP (28) to obtain clean data, and the host sequence was aligned with Bowtie2 (29) to remove reads derived from the host. MEGAHIT (30) assembled high-quality short reads from each DNA sample. MetaGeneMark (31) was used to perform metagenomic gene prediction on the assembled scaffold. CD-HIT (32) was used to cluster predicted genes, and redundant sequences were removed to construct the gene catalog. Salmon (33) was used for quantification. Diamond’s (34) BLASTP function was used for functional annotation, and Kraken2’s default parameters were used for taxonomic annotation.

### LC/MS Nontargeted Metabolomics Analysis

Metabolite extraction was performed according to a previously reported method. In short, samples were extracted by directly adding precooled methanol and acetonitrile (2:1, v/v), and internal standards mix 1 (IS1) and internal standards mix 2 (IS2) were added for quality control of the sample preparation. After vortexing for 1 minute and incubating at −20 °C for 2 hours, the samples were centrifuged for 20 minutes at 4000 rpm, and the supernatant was then transferred for vacuum freeze drying. The metabolites were resuspended in 150 µL of 50% methanol and centrifuged for 30 min at 4000 rpm, and the supernatants were transferred to autosampler vials for LC‒MS analysis. A quality control (QC) sample was prepared by pooling the same volume of each sample to evaluate the reproducibility of the whole LC‒MS analysis. For metabolite separation and detection, a Waters 2D UPLC (Waters, USA) tandem Q Exactive high-resolution mass spectrometer (Thermo Fisher Scientific, USA) was used. The samples were analyzed using a Waters 2D UPLC (Waters, USA) coupled to a Q-Exactive mass spectrometer (Thermo Fisher Scientific, USA) with a heated electrospray ionization (HESI) source and it was controlled by the Xcalibur 2.3 software program (Thermo Fisher Scientific, Waltham, MA, USA). The separation was carried out on a Waters ACQUITY UPLC BEH C18 column (1.7 m, 2.1 mm, 100 mm, Waters, USA), with the column temperature set to 45 °C. In positive mode, the mobile phase contained 0.1% formic acid (A) and acetonitrile (B), while in negative mode, the mobile phase contained 10 mM ammonium formate (A) and acetonitrile (B). The gradient conditions were as follows: 0–1 min, 2% B; 1–9 min, 2%–98% B; 9–12 min, 98% B; and 12.1–15 min, 2% B. The injection volume was 5 µL, and the flow rate was 0.35 mL/min. Compound Discoverer 3.1 (Thermo Fisher Scientific, USA) software was used to process the LC‒MS/MS data, which included peak extraction, peak alignment, and compound identification (35, 36).

### Statistical Analysis

To compare the differences between different microorganisms, the Wilcoxon rank-sum test, Kruskal‒Wallis (KW), LEfSe analysis, Adonis (37), and ANOSIM (38) were used. The metabolomics R package metaX (39) and the metabolome bioinformatic analysis pipeline were used for data preprocessing, statistical analysis, metabolite classification annotations, and functional annotations. To compare metabolites between groups, principal component analysis (PCA), partial least squares discriminant analysis (PLS-DA), Student’s t test, and fold change obtained from the variability analysis were used. The correlation between microorganisms and metabolites was evaluated using Spearman correlation and random forest analyses. Pearson correlation analysis was used to evaluate the correlation of species and metabolites with the clinical data. The statistical significance level was set at p<0.05.

## RESULTS

After a rigorous pathological diagnosis and exclusion process, 68 patients with hepatocellular carcinoma (HCC), 33 patients with liver cirrhosis (LC), and 34 healthy controls (NC) were included in the analysis and comparison. We performed metagenome sequencing and untargeted LC‒MS analysis on the feces and serum of the participants, respectively. Kruskal‒Wallis, the Wilcoxon rank-sum test, and abundance restriction were used to screen different subgroups of microbes, and LEfSe analysis was used to screen HCC-associated species-level key gut microbiota (KGM). In addition, a metabolic pathway enrichment analysis was performed on the differential metabolites that were screened based on the untargeted LC‒MS results to screen for HCC-related key serum metabolites (KSM) in important metabolic pathways. Using random forest classification models, the potential of various panels consisting of different species or metabolites as biomarkers was evaluated. Then, in fecal- and serum-matched enrollees, a combined analysis of differential species and differential metabolites was performed to determine which omics best separated HCC from non-HCC by comparing separate and combined modeling of different omics with the receiver operating characteristic (ROC) curves. Finally, the best panel’s worth was determined by comparing the difference between the best panel and the best omics model (Figure 1).

**Figure 1.**
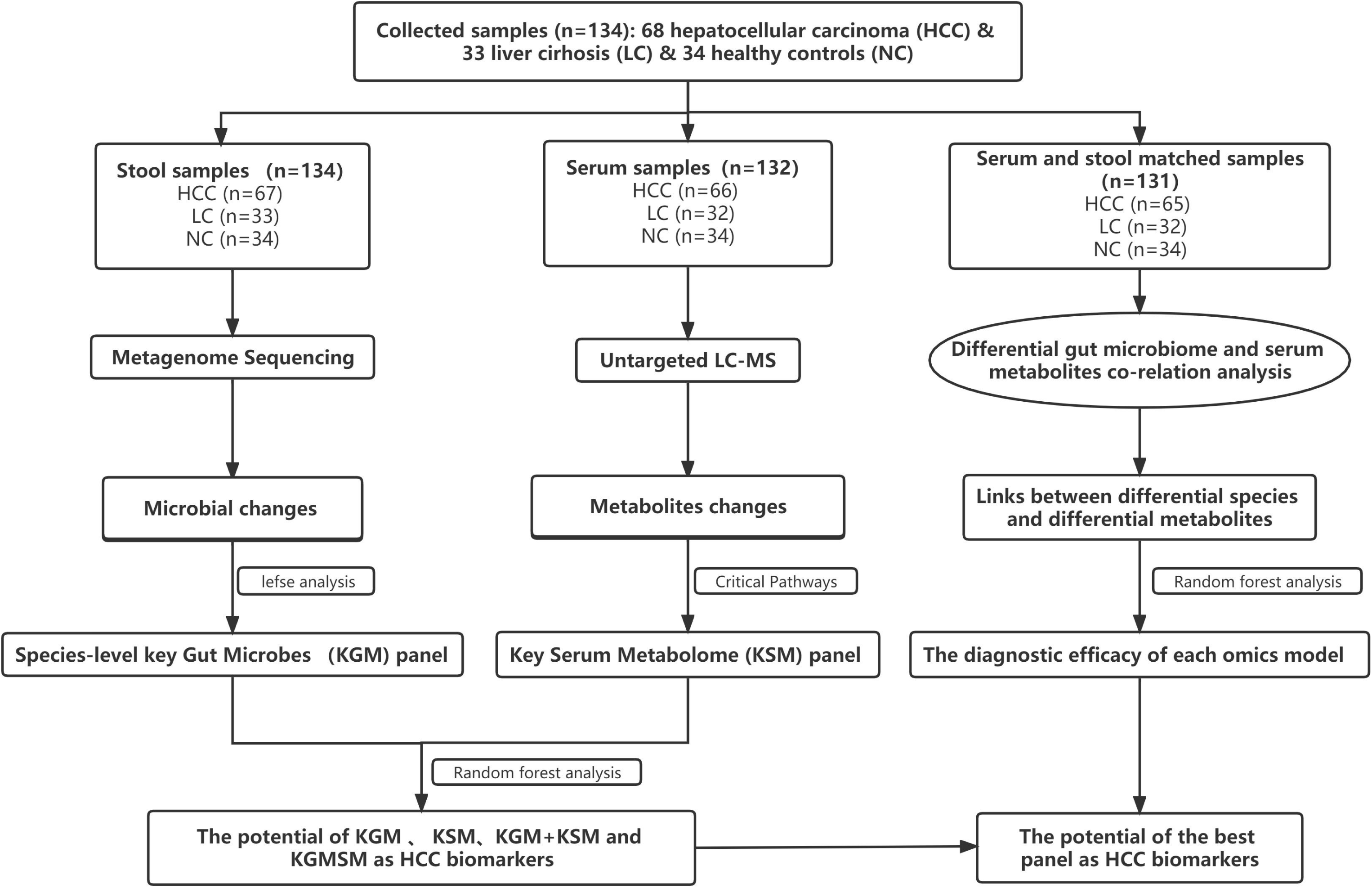
A schematic of the design and the experimental flow diagram. After a strict pathological diagnosis and exclusion process, 68 patients with HCC, 33 patients with cirrhosis, and 34 healthy controls were included at the Second Hospital of Nanjing, Jiangsu Province, China. In total, 132 serum samples and 134 feces samples were included in the analysis. We characterized the gut microbiome of 67 patients with HCC, 33 patients with cirrhosis, and 34 healthy controls and identified the microbial markers. To identify metabolite markers, we simultaneously characterized the serum metabolites from 66 hepatocellular carcinomas, 32 cases of hepatic sclerosis, and 34 healthy controls. We used random forest analysis to assess the ability of various marker combinations to distinguish the HCC cohort from the non-HCC cohort (cirrhosis and healthy controls). We examined the link between the gut microbiota and serum metabolites that changed significantly in HCC using serum- and fecal-matched cohorts. HCC, hepatocellular carcinoma.

### Demographics of the study cohort and quality control of the samples

Before the experimental design and sample collection, participants were matched for dietary habits and clinical characteristics (including age, sex, and body mass index) to ensure that established confounding factors did not affect group differentiation. Serum levels of alanine aminotransferase (ALT), aspartate aminotransferase (AST), glutamyl transpeptidase (GGT), and total bilirubin were significantly higher in HCC patients than in controls, but total protein and platelets were significantly lower (Table 1).

**Table 1.**
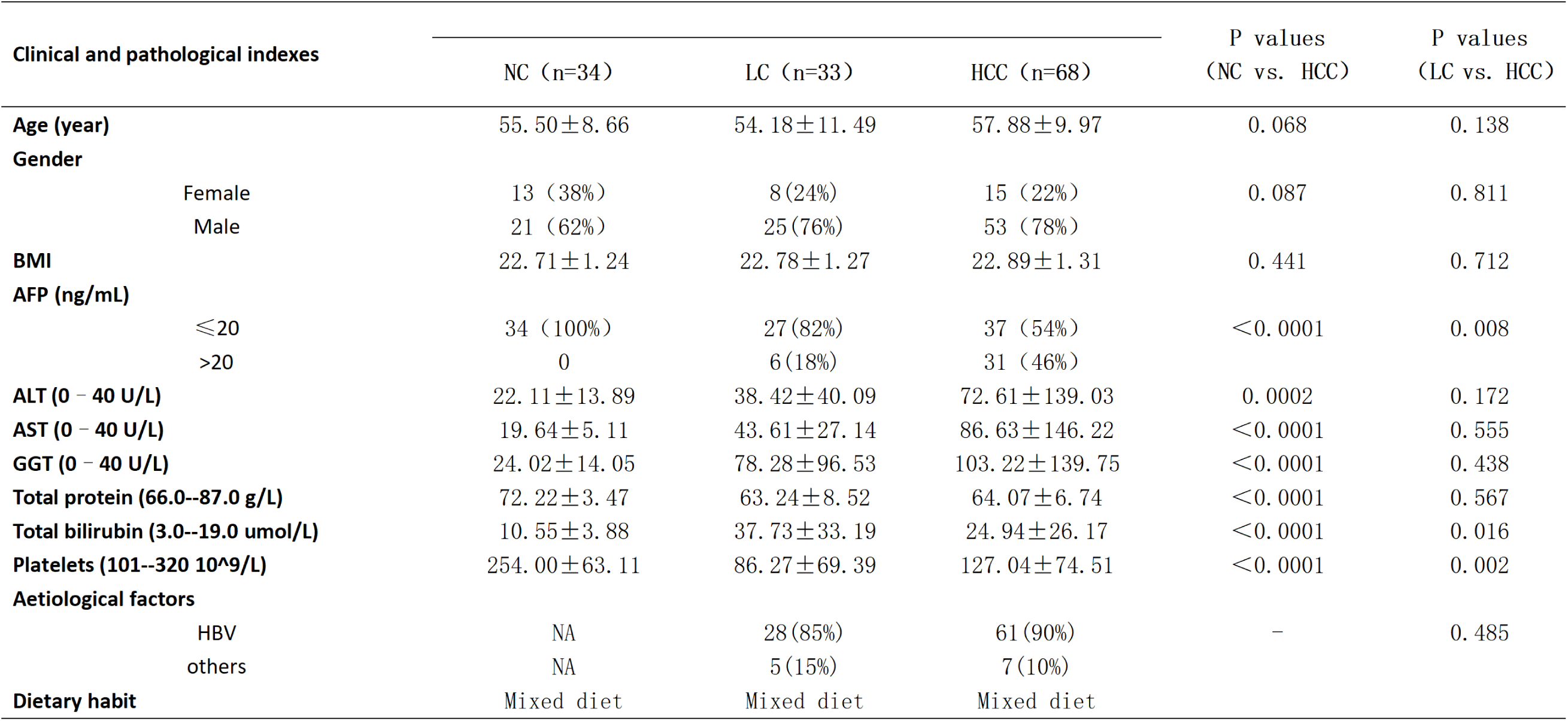
One-way analysis of variance was used to evaluate the differences among the three groups. The Wilcoxon rank-sum test was used to compare the variables between the two groups. BMI, body mass index; AFP, alpha-fetoprotein; ALT, alanine aminotransferase; AST, aspartate aminotransferase; GGT, glutamyl transpeptidase; HBV, hepatitis B virus; HCC, hepatocellular carcinoma; LC, liver cirrhosis; NC, healthy controls; NA, not applicable

| Clinical and pathological indexes |  | NC (n=34) | LC (n=33) | HCC (n=68) | P values<br>(NC vs. HCC) | P values<br>(LC vs. HCC) |
| --- | --- | --- | --- | --- | --- | --- |
| **Age (year)** |  | 55.50±8.66 | 54.18±11.49 | 57.88±9.97 | 0.068 | 0.138 |
| **Gender** |  |  |  |  |  |  |
|  | Female | 13 (38%) | 8 (24%) | 15 (22%) | 0.087 | 0.811 |
|  | Male | 21 (62%) | 25 (76%) | 53 (78%) |  |  |
| **BMI** |  | 22.71±1.24 | 22.78±1.27 | 22.89±1.31 | 0.441 | 0.712 |
| **AFP (ng/mL)** |  |  |  |  |  |  |
|  | ≤20 | 34 (100%) | 27 (82%) | 37 (54%) | <0.0001 | 0.008 |
|  | >20 | 0 | 6 (18%) | 31 (46%) |  |  |
| **ALT (0 – 40 U/L)** |  | 22.11±13.89 | 38.42±40.09 | 72.61±139.03 | 0.0002 | 0.172 |
| **AST (0 – 40 U/L)** |  | 19.64±5.11 | 43.61±27.14 | 86.63±146.22 | <0.0001 | 0.555 |
| **GGT (0 – 40 U/L)** |  | 24.02±14.05 | 78.28±96.53 | 103.22±139.75 | <0.0001 | 0.438 |
| **Total protein (66.0--87.0 g/L)** |  | 72.22±3.47 | 63.24±8.52 | 64.07±6.74 | <0.0001 | 0.567 |
| **Total bilirubin (3.0--19.0 umol/L)** |  | 10.55±3.88 | 37.73±33.19 | 24.94±26.17 | <0.0001 | 0.016 |
| **Platelets (101--320 10<sup>9</sup>/L)** |  | 254.00±63.11 | 86.27±69.39 | 127.04±74.51 | <0.0001 | 0.002 |
| **Aetiological factors** |  |  |  |  |  |  |
|  | HBV | NA | 28 (85%) | 61 (90%) | – | 0.485 |
|  | others | NA | 5 (15%) | 7 (10%) |  |  |
| **Dietary habit** |  | Mixed diet | Mixed diet | Mixed diet |  |  |

Stool morphology and color were assessed prior to metagenome sequencing. Except for one stool sample with an abnormal color in the HCC group, all stool samples were yellow and soft, with no significant differences among the groups (online Supplementary Table S2). To ensure the accuracy of the subsequent analysis, the raw sequencing data from 134 stools (67 HCC, 33 LC, and 34 NC) were filtered and assembled for statistical analysis and gene prediction (online Supplementary Table S3).

After the removal of hemolyzed serum samples, a total of 132 serum samples (66 HCC, 32 LC, and 34 NC) were included in the analysis (online Supplementary Table S1). The base peak chromatograms (BPC) of all QC samples overlapped, the spectrum overlap was good, and the retention time and peak response intensity fluctuated very little, indicating that the instrument was in good condition and that the signal was stable throughout the entire sample detection and analysis (supplementary online figure S2A). The ratio of compounds in the QC sample with a relative peak area CV of less than or equal to 30% to the total number of compounds was higher than 60%, indicating that the data quality was sufficient (online supplementary figure S2B).

### Intestinal flora structural changes in hepatocellular carcinoma

We conducted statistical analyses of microbial abundance for each of the three groups. The estimated species richness in each group was close to saturation according to the rarefaction analysis (Figure 2A). The Chao1 indices of the different groups differed significantly at both the phylum and genus levels, whereas the Shannon and Simpson indices differed significantly at the genus level but not at the phylum level (online Supplementary Figure S3). At the species level (online Supplementary Table S4), the Chao1 indices revealed that the community richness differed significantly among the groups, with NC > HCC > LC (Figure 2B); Shannon indices and Simpson indices demonstrated that the microbiome diversity of HCC and LC was significantly lower than that of NC, with the diversity of HCC being higher than that of LC (with no significance) (Figure 2C-D).

**Figure 2.**
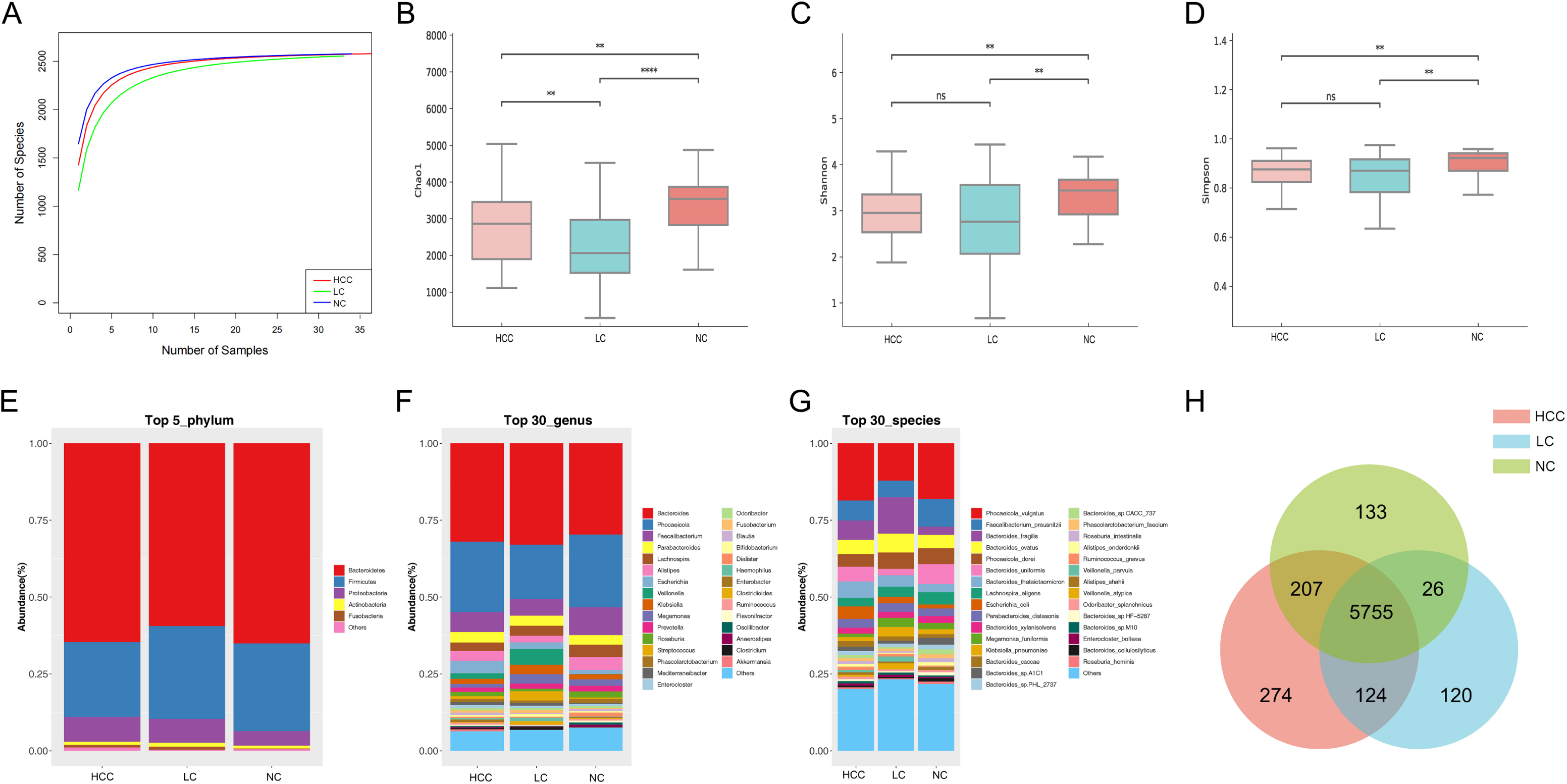
The gut microbiome community is divided into three groups. (A) Rarefaction curves between the number of samples and the number of species. In all samples, the number of species approached saturation. Fecal microbial alpha diversity at the species level was estimated by the Chao1 index (B), Shannon index (C), and Simpson index (D). * p < 0.05, ** p < 0.01, *** p < 0.001 (E) The top five representative phyla and their proportions in each of the three groups. (E) The top 5 representative phyla and their proportions among the three groups. (F-G) The top 30 representative genera and species, as well as their proportions in each of the three groups. (H) A Venn diagram displaying group overlaps revealed that 274 of the total richness of 6640 species were unique to HCC. The red circle represents HCC, the blue circle represents LC, and the green circle represents NC.

We investigated the structure of the intestinal flora communities of the three groups (online Supplementary Table S5). Bacteroidetes, Firmicutes, and Proteobacteria accounted for more than 90% of the total abundance and were the dominant phyla in the three groups (Figure 2E). At the genus level, Bacteroides and Phocaeicola were the most dominant in all three groups, with the relative abundance of Bacteroides increasing and Phocaeicola decreasing in HCC and LC compared to NC (Figure 2F). At the species level, except for *Phocaeicola vulgatus*, which had the highest abundance in all three groups, the most abundant species in HCC were *Faecalibacterium prausnitzii* (6.48%), *Bacteroides fragilis* (6.32%), and *Bacteroides thetaiotaomicron* (5.35%), and in LC, they were *Bacteroides fragilis* (11.8%), *Bacteroides ovatus* (6.14%), and *Faecalibacterium prausnitzii* (5.41%). Additionally, the most prevalent bacteria in NC were *Faecalibacterium prausnitzii* (8.99%), *Bacteroides uniformis* (6.51%), and *Phocaeicola dorei* (5.16%) (Figure 2G). Additionally, Venn plots of the intergroup overlap showed that 5755 of the 6640 species found were shared by all three groups, while 274 species were found only in HCC (Figure 2H).

### Differential analysis of intestinal microbes

To compare the differences in fecal microbial communities between groups and to identify microbes associated with HCC, Kruskal‒Wallis was performed on HCC, LC, and NC, and microbes with a p value<0.05 and median relative abundance greater than 0.01% of the total abundance were recognized as differential microbes. The results showed that a total of 4 phyla, 49 genera, and 86 species were identified (p < 0.05) (online Supplementary Table S6). The Wilcoxon rank-sum test was used to compare the differences in microbes among the groups (online supplementary figure S4A–C).

At the phylum level, the number of Verrucomicrobia was significantly higher in HCC patients than in LC patients, whereas Cyanobacteria, Euryarchaeota, and Uroviricota were lower in HCC patients than in NC patients (p<0.05) (online Supplementary Table S7). At the genus level, 21 genera, including Roseburia, Lachnospira, and Ruminococcus, were significantly higher in HCC than LC, while 42 genera, excluding Veillonella, were significantly lower in HCC than NC, including Faecalibacterium, Alistipes, and Phaecolarctobacterium. (online Supplementary Table S8). Correspondingly, 35 species, such as *Phocaeicola vulgatus*, *Lachnospira eligens*, *Bacteroides uniformis*, and *Ruminococcus bicirculans*, differed between HCC and LC (p<0.05). Compared to NC, except for *Veillonella parvula*, *Veillonella* sp. T1–7, *Veillonella atypica*, and *Veillonella dispar*, which were significantly increased in HCC, all 57 species (*Phocaeicola dorei*, *Bacteroides uniformis*, *Faecalibacterium prausnitzii*, etc.) were significantly reduced (P<0.05) (online Supplementary Table S9). Furthermore, we compared the bacterial differences between LC and NC at the phylum and species levels, and the results are attached (online Supplementary Table S7-9).

To identify the key gut microbiota, we used linear discriminant analysis (LDA) and its effect size (LEfSe). After excluding species with relative abundances of less than 0.01%, the HCC, LC, and NC groups contained 2, 9, and 30 species, respectively (Figure 3). *Odoribacter splanchnicus* and *Ruminococcus bicirculans* were species-level potential biomarkers for the detection of HCC.

**Figure 3.**
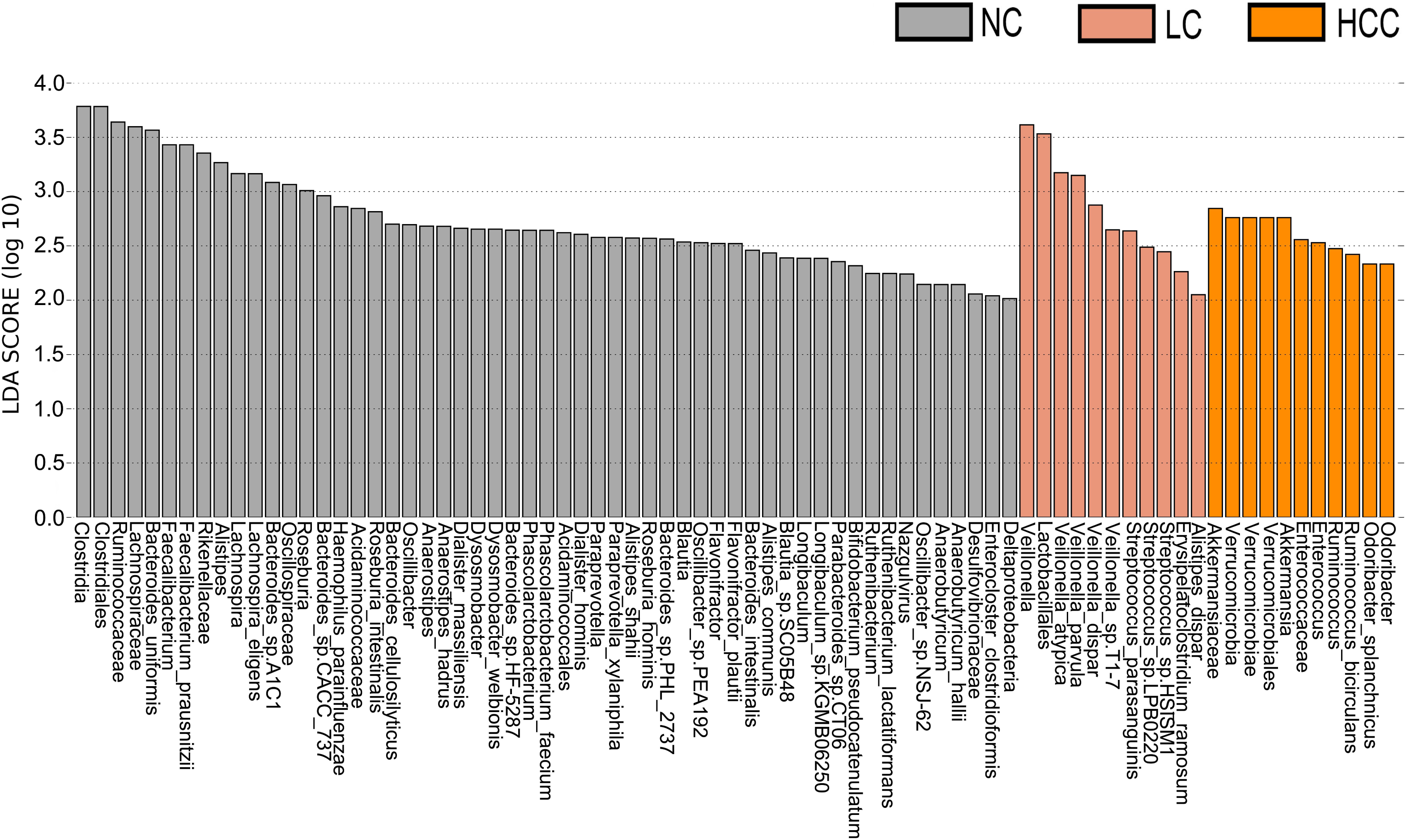
Key species selection by linear discriminant analysis (LDA) effect size (LEfSe). Differential microbial score chart: the higher the score, the greater the contribution of the microbe to the difference.

### Serum metabolite changes in patients with hepatocellular carcinoma

Tumorigenesis is accompanied by a general change in the metabolite status of the local tissue and circulatory system. Metabolites and fermentation products produced by the intestinal flora can enter the bloodstream and impact the host’s physiological functions. The metabolic profile in the serum was examined to investigate the relationship between metabolites in the serum and HCC. We discovered 8,709 compounds, 2,934 of which were identifiable (online Supplementary Table S10). Based on the abundance of metabolites detected by untargeted metabolomics, partial least squares-discriminant analysis (PLS-DA) was performed (Figure 4A–B). According to the scatter plot, the samples from HCC, LC, and NC were separable, and the alignment test revealed that the data were not overfitted (Figure 4C–D) (in general, the closer the slopes of the R2Y and Q2Y lines are to zero, the more likely the model is overfitted). The PLS-DA analysis for LC versus NC is depicted in online supplementary figure S5A-B.

**Figure 4.**
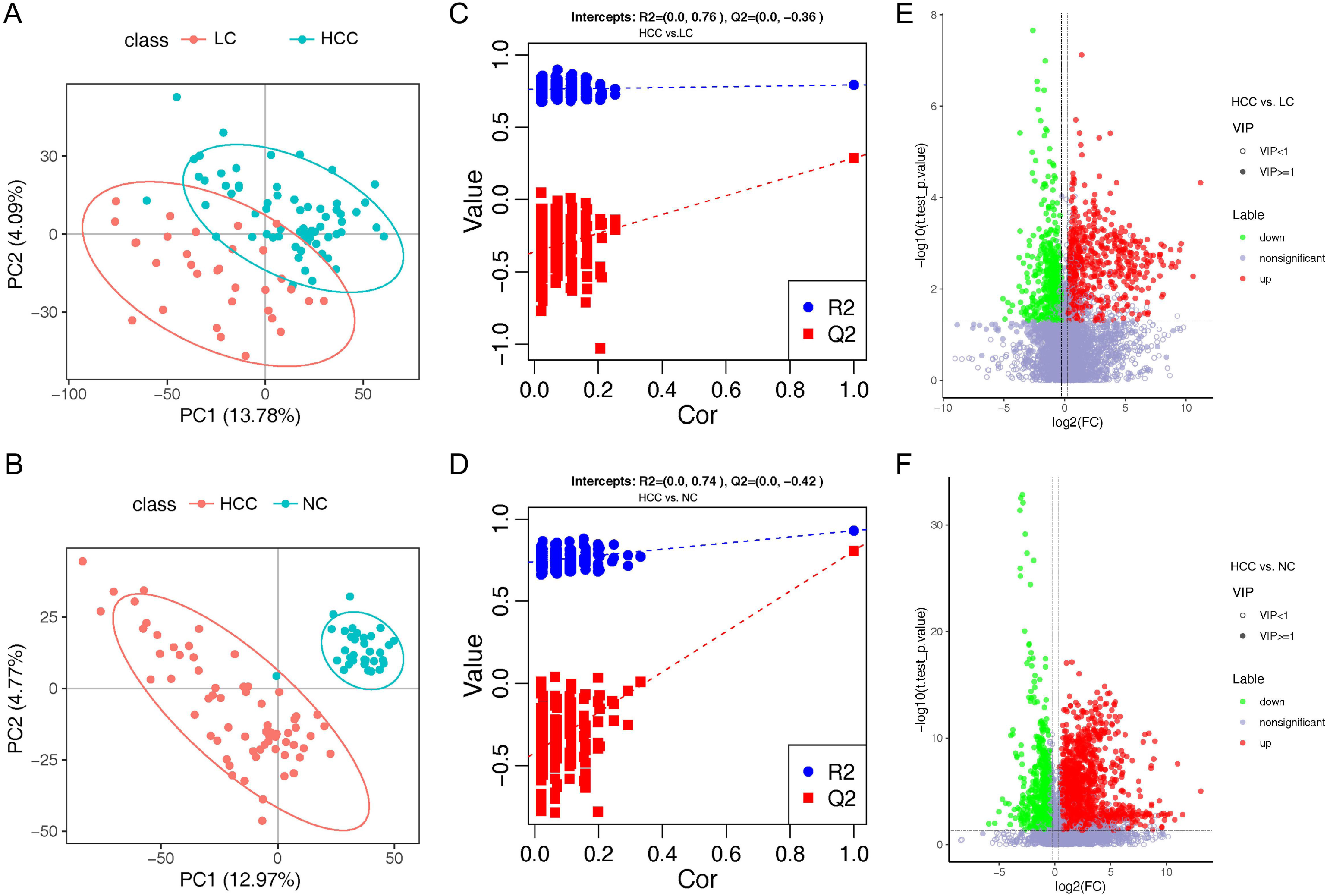
Overview of altered serum metabolism in HCC (n=66) and non-HCC (LC (n=32), NC (n=34)). (A-B) PLS-DA shows the differences between the groups’ metabolites. The abscissa (PC1) and the ordinate (PC2) are the two main coordinates that explain the greatest difference between the samples. The number is the score of the principal component, which represents the percentage of the explanation on the overall variance of the specific principal component. The graph points represent samples, and different colors represent various sample grouping information; similar samples are clustered together. (C-D) The two rightmost points in the figure are the actual R2Y and Q2 values of the model, and the remaining points are the R2Y and Q2 values obtained by randomly arranging the samples used. This result is mainly used to judge whether the model is overfitting and the validity of the model. A volcano plot is a graphical representation of differential metabolism. (E) Metabolites that differ between HCC and NC. (F) Metabolites that differ between HCC and LC. Green marks the downregulated differential metabolites, red marks the upregulated differential metabolites, and metabolites without differences are labeled purple‒gray.

PLS-DA yielded “Variable Important for the Projection” (VIP) values, with larger values indicating a greater contribution of the variable to the subgroup. The following criteria were used to screen biologically significant differential metabolites: 1) VIP value≥1 for the PLS-DA’s first principal component, 2) p value<0.05 for the t test, and 3) fold-change≥1.2 or ≤0.83. In HCC versus LC, HCC versus NC, and LC versus NC, 424, 823, and 825 differential metabolites were screened for biological significance, respectively (online Supplementary Table S11). The differences in metabolism between HCC and non-HCC were demonstrated using volcano plots (Figure 4E–F). The volcano plots of LC versus NC are shown in online supplementary figure S5C.

To better understand the mechanism of differential metabolites implicated in the pathogenesis of HCC, we performed metabolic pathway enrichment analysis on the KEGG IDs of the differential metabolites. We considered metabolic pathways with p values less than 0.05 to be significantly enriched in differential metabolites and plotted bubble plots (Figure 5A-C) for these pathways (the metabolites on the pathways are shown in online Supplementary Table S12). Bile secretion, cholesterol metabolism, purine metabolism, caffeine metabolism, metabolic pathways, apoptosis, and vitamin digestion and absorption were all significantly enriched in the HCC versus LC comparison group. The pathways that were significantly enriched in the HCC versus NC comparison group included caffeine metabolism, metabolic pathways, bile secretion, cholesterol metabolism, primary bile acid biosynthesis, drug metabolism - other enzymes, prostate cancer, and porphyrin and chlorophyll metabolism. In the LC versus NC comparison group, the differential metabolites were involved in caffeine metabolism, bile secretion, cholesterol metabolism, primary bile acid biosynthesis, metabolic pathways, porphyrin and chlorophyll metabolism, and cysteine and methionine metabolism.

**Figure 5.**
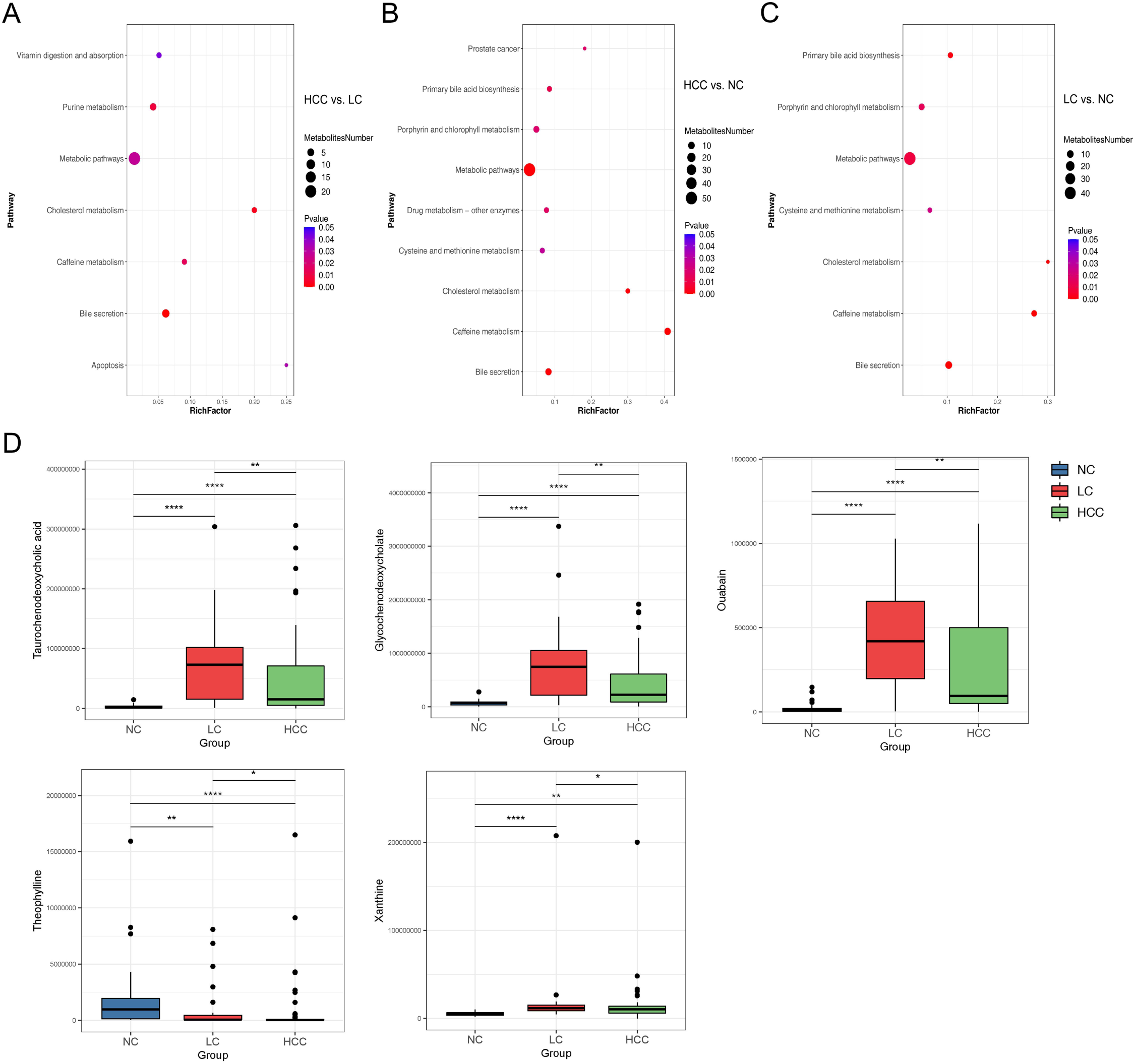
(A-C) Bubble plots of pathways with significant enrichment of differential metabolites. The ordinate is the name of the metabolic pathway, and the abscissa is the rich factor (rich factor = the number of differential metabolites annotated to the pathway/all identified metabolites annotated to the pathway). The larger the rich factor, the greater the proportion of differential metabolites annotated to the pathway. The color from blue to red indicates that the p value decreases sequentially; the larger the point, the more differential metabolites are enriched in the pathway. (D-H) Distribution of different metabolites in each group. *p < 0.05, **p < 0.01, ***p < 0.001.

We discovered that the bile secretion, cholesterol metabolism, and caffeine metabolism pathways are all associated with the progression of HCC. We focused on three pathway-related differential metabolites involved in all three comparison groups to find metabolomic markers: ouabain, taurochenodeoxycholic acid (TCDCA), glycochenodeoxycholate (GCDCA), theophylline, and xanthine (online Supplementary Table S13). We researched the expression of these metabolites in various groups (Figure 5D–H). The findings revealed that the expression of ouabain, TCDCA, and GCDCA was significantly lower in HCC than in LC but significantly higher than in NC. Theophylline expression was significantly lower in HCC compared to non-HCC, whereas xanthine expression was significantly higher in HCC compared to NC.

### Correlation analysis of metagenome and metabolome

We performed correlation analysis and joint analysis of serum and fecal matched microbiome-metabolome data (65 in HCC, 32 in LC, and 34 in NC) to explore the relationship between the microbiota and the serum metabolome. We calculated Spearman correlation coefficients between different species and metabolites and obtained a correlation coefficient matrix (online Supplementary Table S14), as well as the top 20 differential species and metabolites with the smallest p values for each omics, which were chosen for the heatmap (Figure 6A-B), and the results of LC versus NC are shown in supplementary figure S6A.

**Figure 6.**
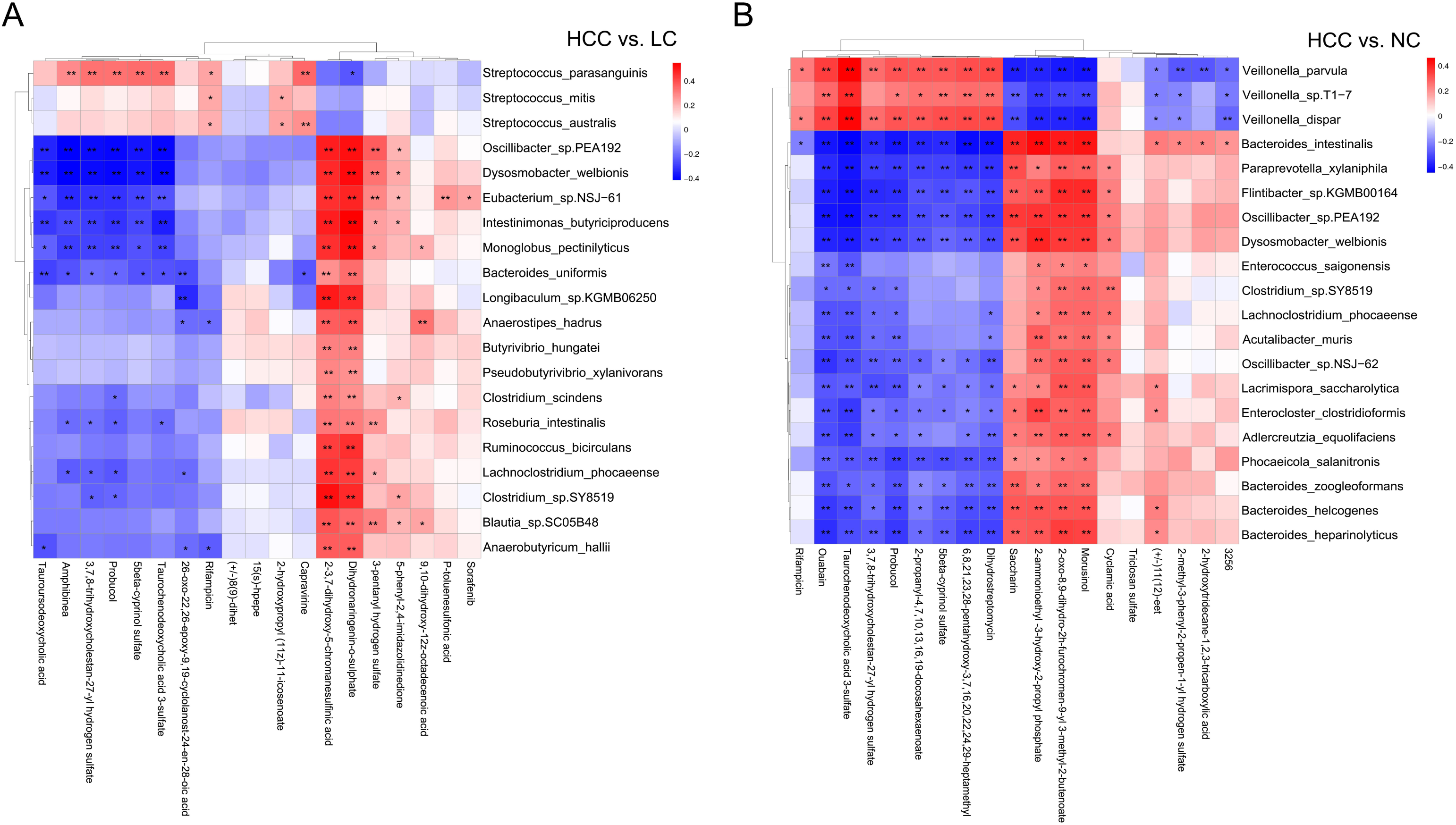
The heatmap of the top 20 differential species and differential metabolites with the smallest p values for every omics in HCC vs. non-HCC. Columns represent the differential metabolites, and rows represent the differential species. The color blocks represent the correlation coefficient. The darker the color, the stronger the correlation between the different species and the different metabolites. Red represents a positive correlation, and blue represents a negative correlation. * represents p<0.05, ** represents p<0.01.

We performed an association analysis of *Odoribacter splanchnicus* and *Ruminococcus bicirculans* with differential metabolites to screen for key species-associated serum metabolites and discovered 66 key species with significantly associated metabolites in HCC versus LC and 45 in HCC versus NC (p < 0.05) (Online Supplementary Table S15). In both comparison groups, nine metabolites showed significant associations with key species associated with HCC, including three key metabolites (TCDCA, GCDCA, and xanthine) associated with HCC (Figure 7A).

**Figure 7.**
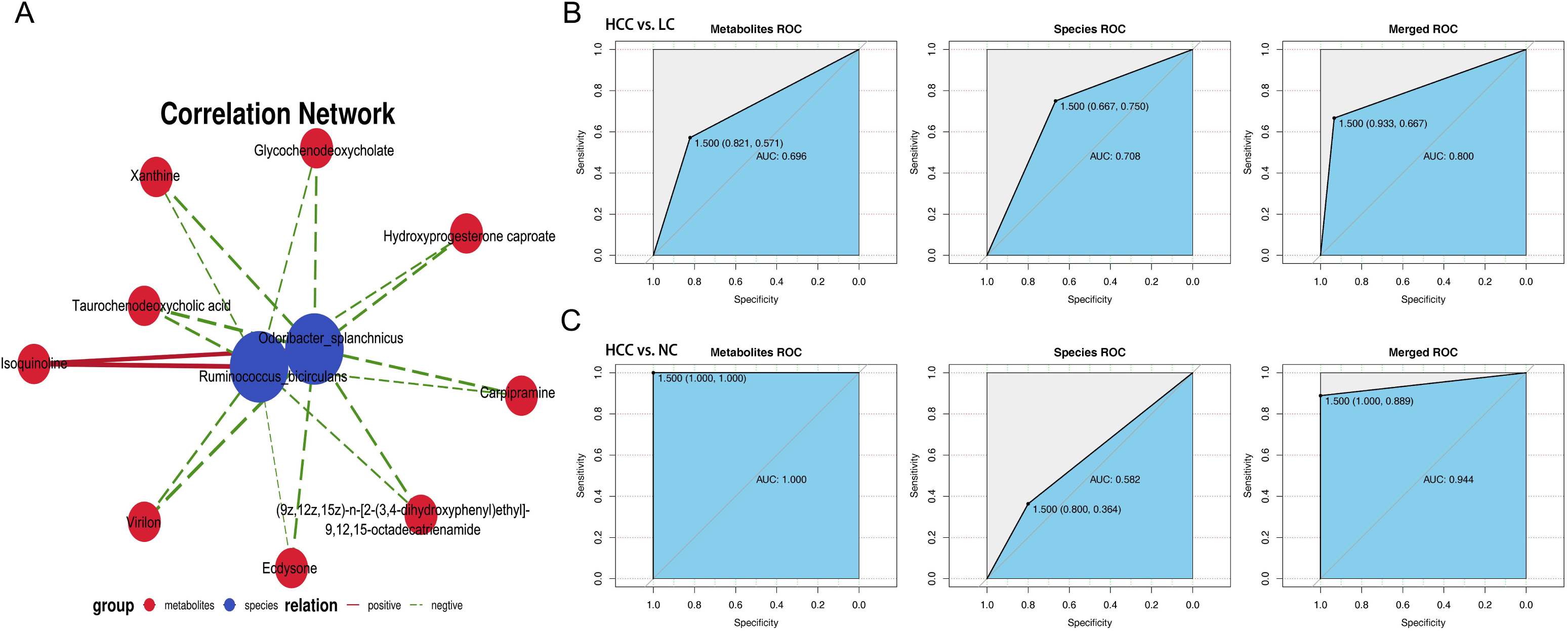
Correlation analysis of the metagenome and metabolome. (A) Spearman correlation network interaction diagram of the key species and differential metabolites. Each point in the figure represents a species or a metabolite. The more lines there are between the points, the more species or metabolites it may regulate. Green dots represent species, red dots represent metabolites, red connecting lines between dots are positive correlations, and green lines are negative correlations. The thickness of the line represents the level of the correlation coefficient. (B-C) The ROC curves of the random forest analysis of species and metabolism biomarkers. Random forest ROC map of species and metabolomes (the ROC map of the metabolome is on the left, the ROC map of species is in the middle, and the ROC map of the species and metabolomes is on the right).

By comparing the ROC curves between the separate modeling of different omics and the combined data modeling, we evaluated which omics better separates HCC and non-HCC, and we discovered that in HCC versus LC, merged > species > metabolites with AUC values of 0.800, 0.708, and 0.696, respectively (Figure 7B). Metabolites > merged > species had AUC values of 1.000, 0.944, and 0.582 for HCC versus NC, respectively (Figure 7C). The results of LC versus NC are shown in Supplementary Figure S6B.

### Evaluation of the contribution of various panels to the prediction of HCC using ROC curves

To evaluate the biomarker potential of HCC-related keystone species and metabolites, we constructed a key gut microbiota (KGM) panel with *Odoribacter splanchnicus* and *Ruminococcus bicirculans* and a key serum metabolite (KSM) panel with ouabain, TCDCA, GCDCA, theophylline, and xanthine. We tested each panel’s ability to distinguish between HCC and non-HCC. Based on the relative abundance of metagenome and untargeted metabolic profile assays, we divided the data from each panel into a training set and a validation set, first building a random forest model for the training set and then using this model to predict the validation set and construct ROC curves. We perform 10-fold cross-validation and then averaged the resulting ROC curve. The AUC values of the KGM panel for HCC versus NC and HCC versus LC were 0.60±0.22 and 0.65±0.19, respectively (Figure 8A), whereas the AUC values of the KSM panel for distinguishing HCC from NC and LC were 0.95±0.06 and 0.65±0.15, respectively (Figure 8B). It is also worth noting that the KSM panel did surprisingly well to distinguish LC from the NC group (AUC: 0.93±0.12) (online supplementary figure S5D). After that, we incorporated the KGM and KSM data into the random forest model to develop the ROC curve. The AUC values of the KGM+KSM panel for HCC versus NC and HCC versus LC were 0.97±0.06 and 0.72±0.18, respectively (Figure 8C).

**Figure 8.**
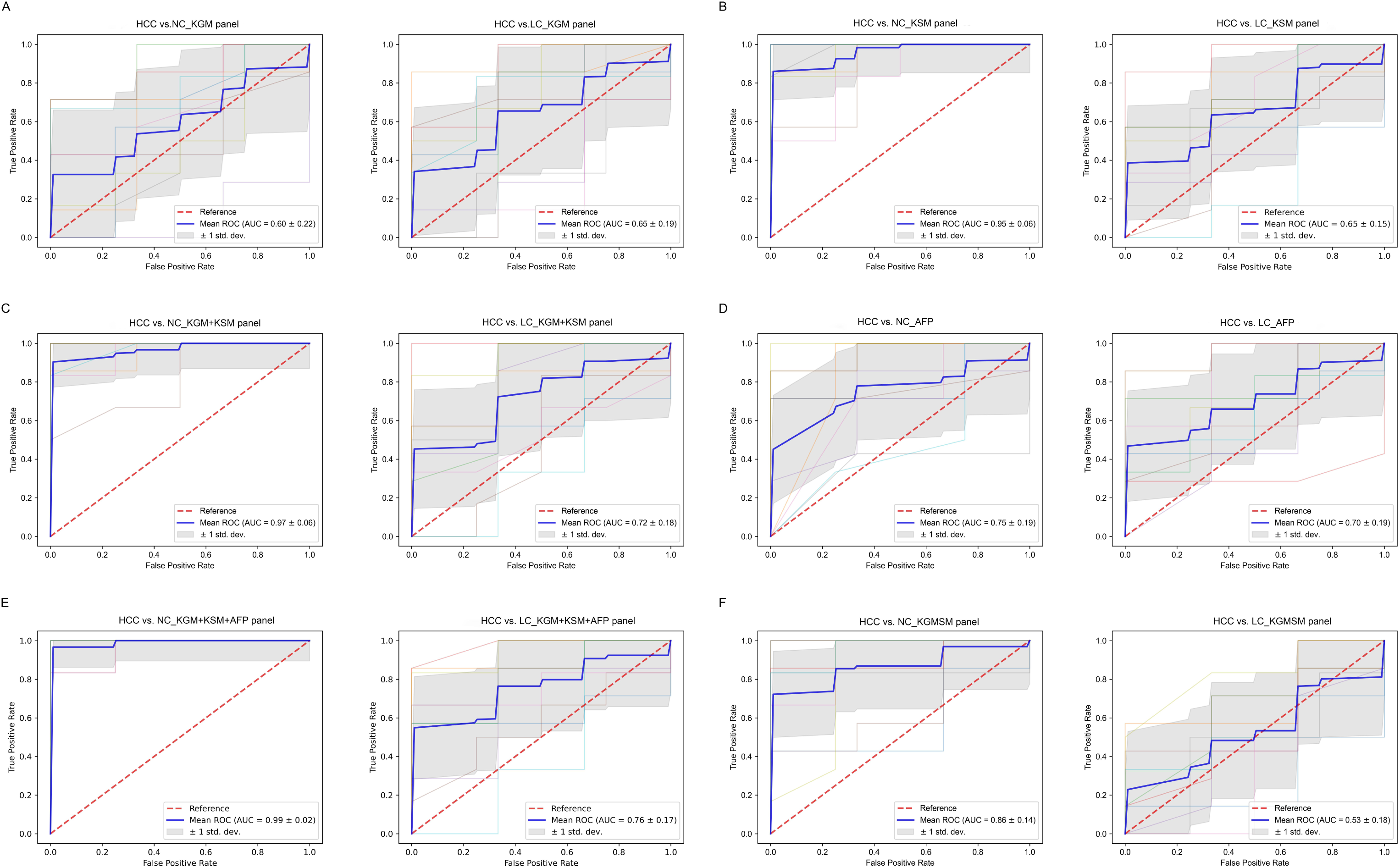
The ROC curves of a random forest analysis of different panels. KGM, key gut microbes; KSM, key serum metabolites; KGMSM, key gut microbial-associated serum metabolites. The abscissa of the ROC curve is the false-positive rate; the ordinate is the true positive rate; the blue curve is the average curve after 10 folds; the AUC is the area under the curve; the shaded region is the upper and lower 1 standard deviation.

The clinical indicator alpha-fetoprotein (AFP) (cutoff value of 20 ng/mL) is commonly used to aid in the diagnosis of HCC. To compare the efficacy of AFP and our KGM+KSM panel in detecting HCC, we recorded AFP levels in all individuals included in this study. According to our findings, the AUC values for the AFP panel for HCC versus NC and LC were 0.75±0.19 and 0.70±0.19, respectively (Figure 8D). In contrast, our KGM+KSM panel performed better than the AFP panel in terms of diagnostic value. When KGM+KSM was combined with AFP to build the ROC curve, the AUC values for distinguishing HCC from NC and LC improved slightly when compared to the KGM+KSM panel, reaching 0.99 ± 0.02 and 0.76 ± 0.17, respectively (Figure 8E). Furthermore, we created a KGMSM panel with 9 differential serum metabolites related to key species, which had significantly lower potential as HCC markers than the KGM+KSM panel, with AUC values of 0.86 ± 0.14 and 0.53 ± 0.18 for HCC versus NC and HCC versus LC, respectively (Figure 8F).

### Correlation of key species, key metabolites, and clinical indicators

We performed Spearman correlation on key metabolites and key species associated with HCC, and the results revealed that ouabain, TCDCA, GCDCA, and xanthine had a significant negative correlation with *Odoribacter splanchnicus* and *Ruminococcus bicirculans* (Figure 9A–B). Meanwhile, we conducted a Pearson correlation analysis of key species and key metabolites with AFP, liver function index, and immune cells (Figure 9C–D). *Odoribacter splanchnicus* was found to be significantly and positively correlated with AFP, white blood cells (WBCs), and leukocytes (LYs); GCDCA and TCDCA were significantly and positively correlated with total bilirubin (TBIL) and gamma-glutamyl transferase (GGT) but significantly and negatively correlated with total protein (TP), platelets (PLTs), and immune cells such as LYs, indicating that HCC-related metabolites were closely related to the deterioration of liver function in HCC patients.

**Figure 9.**
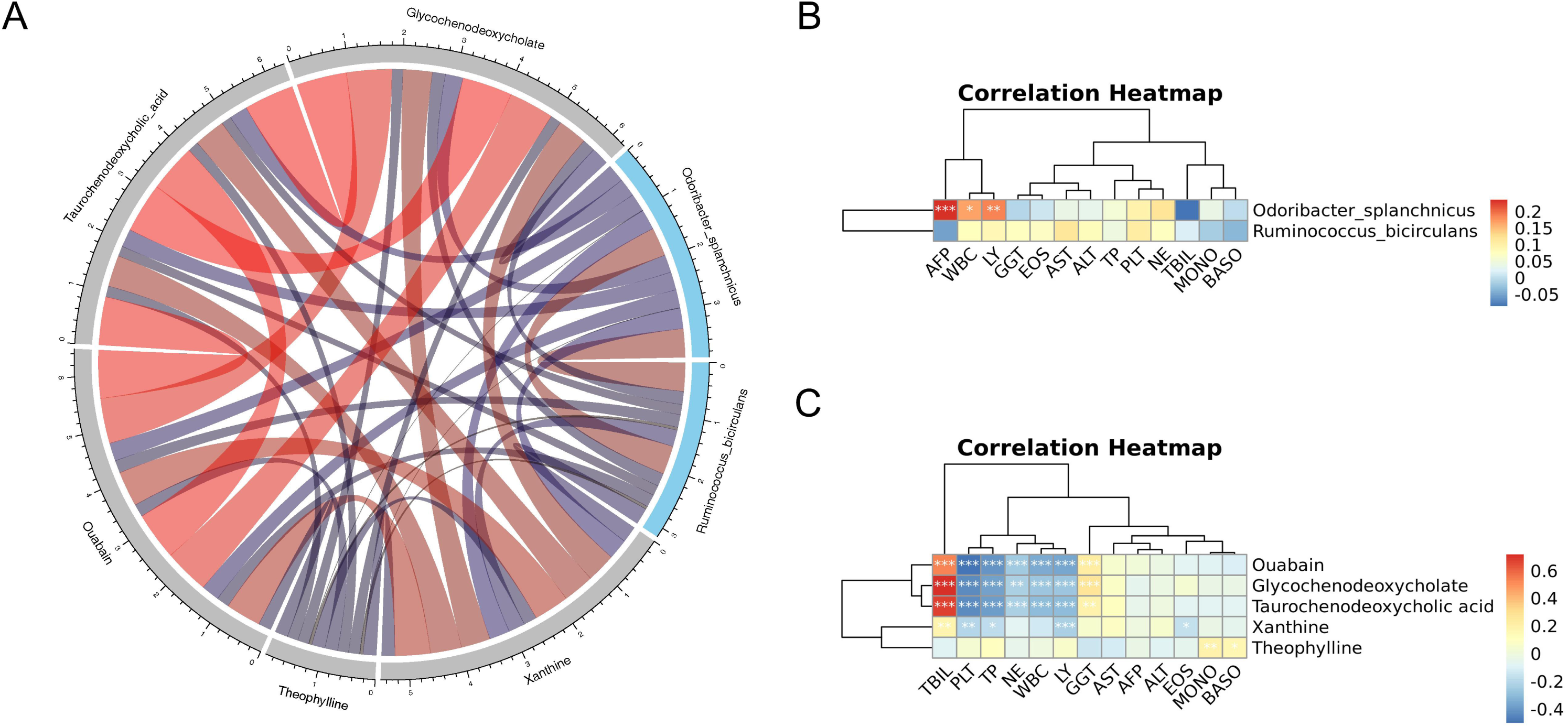
(A) Spearman correlation chord diagram of key species and key metabolites. Species or metabolites are on the edge of the circle in the figure, and the connecting line in the circle represents the correlation between the species and metabolites; red is a positive correlation, and blue is a negative correlation. The darker the color or the thicker the line, the stronger the correlation. (C-D) Pearson correlation cluster heatmap depicting the relationships between the key metabolites (ouabain, TTCDCA, GCDCA, theophylline, and xanthine), the key species (*Odoribacter splanchnicus* and *Ruminococcus bicirculans*) and the clinical indicators. WBC, white blood cell; LY, leukocytes; GGT, gamma-glutamyl transferase; EOS, eosinophil; TP, total protein; PLT, platelets; NE, neutrophilic granulocyte; TBIL, total bilirubin; MONO, monocytes; BASO, basophil. Red indicates positive correlations, whereas blue indicates negative correlations. *p < 0.05, **p < 0.01, ***p < 0.001

## DISCUSSION

Our findings confirmed changes in gut microbiota and serum metabolites in HCC patients. We created panels of key species and key metabolites associated with HCC as potential markers of HCC, both individually and in combination, and then tested the diagnostic value of each panel for HCC. We also compared the two omics to assess the diagnostic performance of different omics alone and in combination. The control group consisted of individuals who underwent a physical examination at Nanjing’s Second Hospital, whereas the HCC patients were newly diagnosed patients. We screened study participants for pharmacological factors such as anticancer drugs and antibiotics to rule out any interference with changes in the intestinal flora. We discovered differences in gut microbial composition and structure among NC, HCC, and LC and discovered that the diversity of fecal microbes decreased in that order, which is consistent with previous research (40).

Fecal Bacteroides are a sign of a healthy gut because they break down polysaccharides and oligosaccharides and give nutrients and vitamins to the host and other intestinal microbes (41). *Bacteroides acidifaciens* in the mouse intestine alleviated liver injury by reducing hepatocyte apoptosis in a cd95-dependent manner, according to one study (42). However, it was discovered in another study on the combined gut microbial and transcriptomic analysis of HCC patients that the extent of tumor load was positively correlated with the abundance of Bacteroides, which caused adverse clinical outcomes via increased serum bile acids (43). Furthermore, Bacteroides has been shown to be a drug target for certain herbal and anticancer drugs in the treatment of HCC, although whether by increasing or decreasing it remains debatable. The Shaoyao Ruangan Mixture, for example, may have antihepatocellular effects by increasing Bacteroides, whereas nimbolide has antihepatocellular effects by decreasing Bacteroides (44, 45). We hypothesize that these disparities are due to differences in animal and human flora, subject selection criteria and heterogeneity, or 16S rRNA sequencing limitations. The detection of species levels is also limited by 16S rRNA sequencing, and while some microbes may maintain dynamic equilibrium at the genus level, species levels within the genus have different effects on disease susceptibility. Our metagenome sequencing results revealed that differential species under the Bacteroides branch, such as Bacteroides_sp. HF-5287, Bacteroides_sp. A1C1, Bacteroides_sp. CACC_737, Bacteroides_sp. PHL_2737, Bacteroides_intestinalis, Bacteroides_uniformis, and Bacteroides_cellulosilyticus were all higher in the NC group than in the other two groups and were also important markers to distinguish the healthy group from the other two groups.

Veillonella was found to be associated with autoimmune hepatitis (AIH), primary biliary cholangitis (PBC), primary sclerosing cholangitis (PSC), HBV infection, and alcoholic hepatitis, among other liver diseases that are highly correlated with liver function indicators and liver inflammation, according to previous research (46, 47). Veillonella was also found to be positively related to AFP, a clinical indicator of HCC (48). This is consistent with our findings that, with the exception of *Veillonella parvula*, *Veillonella sp*. T1–7, *Veillonella atypica*, *Veillonella dispar*, and most key species differing in HCC and LC had a decreasing trend compared to NC. We also discovered that these four species were more abundant in LC than in HCC, which is consistent with the findings of a study on the gut flora of HBV-associated early HCC and LC (49). It is worth noting that the LEfSe analysis results indicate that these four species have the potential to diagnose LC.

Furthermore, *Ruminococcus bicirculans* and *Odoribacter splanchnicus* were identified as potential species-level microbial markers for the diagnosis of HCC in the LEfSe analysis. Ruminococcus’s benefits and drawbacks are debatable. It has been proposed that it is a probiotic that could benefit HCC patients receiving anti-PD-1 immunotherapy as well as combat the anxiety and fear associated with cancer treatment and recurrence (50, 51). Ruminococcus, on the other hand, was strongly associated with some diseases and significantly enriched in patients with thyroid cancer, endometrial cancer, and clear cell renal cell carcinoma, and it may serve as a biomarker for clinical features and prognosis and provide a new therapeutic target for clinical treatment (52, 53, 54). We discovered that the relative abundances of Ruminococcus and *Ruminococcus bicirculans* in HCC were significantly lower than those in NC but significantly higher than those in LC.

In contrast, *Odoribacter splanchnicus*, another potential HCC marker, is widely regarded as a probiotic that can be used in fecal transplantation to treat disease (55). *Odoribacter splanchnicus* was found to be age-enriched in centenarians, and it may aid in health maintenance (56). *Odoribacter splanchnicus* was shown to be useful in treating colitis and colorectal cancer by stimulating IL-6 and IL-1 production and Th17 cell expansion (57). *Odoribacter splanchnicus* was found in this study to be significantly lower in HCC and LC than in NC, and it is expected to be a new therapeutic target for HCC.

Metabolic pathway enrichment analysis was performed using differential metabolites to learn the mechanisms of the metabolic pathway changes in the different groups. Bile secretion, cholesterol metabolism, and caffeine metabolism pathways were found to be enriched in HCC versus LC, HCC versus NC, and LC versus NC, which are closely related to HCC progression. Among the various endogenous metabolites from the host intestinal flora that are synergistically metabolized, bile acids have received increased attention due to their known pro-tumorigenic properties (58, 59), which involve two important receptors: the farnesoid X receptor (FXR) and the G-protein-coupled bile acid receptor (TGR5) (60). There is accumulating evidence that bile acids play an important role in HCC. TCDCA and GCDCA were discovered to be involved in the bile secretion and cholesterol metabolism pathways, and their serum concentrations were both significantly different in pairwise comparisons, suggesting that they could be used as clinical biomarkers. Previous research has discovered that TCDCA not only causes oxidative stress in gastrointestinal tumors, resulting in compensatory upregulation of TR mRNA (61), but it also reduces expression of the tumor suppressor gene CEBP in HepG2 cell lines (62), which is correlated with the risk of colon cancer and HCC (63, 64).

GCDCA is a significant human bile salt. GCDCA treatment of HepG2 cell lines activates ERK1 and ERK2, induces phosphorylation of Mcl-1 at the T163 site and is a potential carcinogen in the development of HCC (65). Meanwhile, in vitro and in vivo studies revealed that GCDCA activated autophagy in HCC cells and significantly increased their invasive potential (66). These mechanisms may represent a novel treatment for HCC. TCDCA and GCDCA were significantly higher in the HCC group than in the NC group in our study.

Another important pathway involving theophylline and xanthine is caffeine metabolism. In the majority of observational studies and meta-analyses, coffee consumption has been linked to a lower risk of cancers such as colorectal (67), breast (68), prostate (69), and liver cancer (70). Theophylline is a xanthine derivative that is primarily eliminated by liver metabolism and is used to treat respiratory diseases such as asthma. Studies have shown that increasing coffee consumption raises serum levels of the metabolite theophylline, which has been shown to have anticancer activity and a protective effect against cisplatin-induced GFR damage in patients with various malignancies, although the precise mechanism is unknown (71, 72). Theophylline is a natural substance that is easily accessible. It has the potential to be modified and used as a scaffold structure for the creation of effective antitumor medications. Non-small cell lung cancer (NSCLC) has been reported to be effectively treated with theophylline derivatives containing 1,2,3-triazole rings (73). According to our findings, theophylline was significantly reduced in patients with HCC.

Additionally, we discovered that 9 serum metabolites, including TCDCA, GCDCA, and xanthine, were closely related to the previously screened key species *Odoribacter splanchnicus* and *Ruminococcus bicirculans* and that the KGMSM panel constructed from the 9 differential metabolites associated with the key species has some diagnostic potential. The Spearman correlation test showed that ouabain, TCDCA, GCDCA, and xanthine all had significant negative correlations with *Odoribacter splanchnicus* and *Ruminococcus bicirculans*.

Previous research has shown that both gut microbes and serum metabolites have great potential for disease diagnosis (40, 74). However, the causal relationship between gut microbes and metabolites in HCC is unclear, and no articles have been published that report on which is best for diagnosing HCC: gut microbes or metabolites. ROC curves were used to assess each panel’s potential contribution to predicting HCC, and we found that the KSM panel was superior to the KGM panel in distinguishing HCC from non-HCC but it had an AUC value of less than 0.7 in distinguishing HCC from LC. When we used the KGM+KSM panel for the diagnosis of HCC, the AUC values for HCC versus LC (AUC > 0.7) and HCC versus NC improved, indicating that the KGM+KSM panel is superior to the KGM panel and the KSM panel as a potential marker for HCC. Using the best ROC results from separate omics modeling versus combined data modeling, the AUCs for HCC versus LC and HCC versus LC were 0.800 (merged) and 1.000 (metabolites), which are very close to the corresponding AUC values of 0.72 and 0.97 for our KGM+KSM panel. Furthermore, when the clinical indicator AFP was included in the panel, the AUC values reached 0.76 and 0.99. Our findings suggest that our KGM+KSM panel could be a promising, noninvasive HCC detection method.

It is worth noting that although the changes and associations of the microbiome and metabolome in HCC were described in our study, which evaluated and compared different panels as HCC markers, these results were not validated in a separate population cohort. In the future, more and larger cohort studies will be needed.

Meanwhile, this study has some limitations. For starters, the prognosis is an important aspect of disease research. We were unable to study the disease’s prognosis for the time being because it was not possible to follow all patients in the short term. Second, the strict enrollment criteria resulted in a small number of patients being enrolled. In the future, we hope to increase the sample size and conduct additional studies through multicenter collaboration. Finally, we focused on two HCC-related key species and five HCC-related key metabolites. We discovered their possible involvement in HCC through pathway enrichment analysis, but this needs to be validated in vivo and in vitro experiments, which is what we will do next. We hope that our research will lead to new approaches to the diagnosis and treatment of HCC.

## CONCLUSIONS

The intestinal flora and serum metabolism in HCC patients were studied. The results imply that HCC could be caused by a mutual regulation of key species and key metabolites. A comparison of the diagnostic performance of the KGM, KSM, and KGM + KSM panels and AFP suggested that HCC-related key species (*Odoribacter splanchnicus* and *Ruminococcus bicirculans*) and key metabolites (ouabain, TCDCA, GCDCA, theophylline, and xanthine) may be potential markers for the diagnosis of hepatocellular carcinoma.

## Data Availability

All data produced in the present study are available upon reasonable request to the authors
All data produced in the present work are contained in the manuscript
All data produced are available online

## ACKNOWLEDGEMENTS

This study was supported by funding for the Infectious Disease Clinical Center of Nanjing and the Infectious Disease Innovation Center of Jiangsu.

## AUTHOR’S CONTRIBUTIONS

X Li conducted all experiments and wrote the manuscript. X Li and X Gu collected samples. Y Yi and J Li revised the manuscript. All authors have read and agreed to the published version of the manuscript.

---

**Supplementary figure S1.** Experimental Workflow and Bioinformatic Analysis Workflow. (A) DNBSEQ library construction and sequencing. We set up strict quality control in each step in experimental workflow (such as sample processing, library prepareation, and sequencing). (B) The bioinformatic analysis workflow for metagenomics. High-quality reads assembly, gene prediction, gene de-redundancy and subsequent abundance construction, as well as species/function differential analysis.

**Supplementary figure S2.** Data Quality Control.(A) BPC overlay of QC samples. BPC is a map that continuously depicts the intensity of the strongest ions in the mass spectrum at each time point.The BPC of all QC samples were overlapped, the spectrum overlap was good, and the retention time and peak response intensity fluctuated little, indicating that the instrument was in good condition and the signal was stable during the whole sample detection and analysis. (B) CV distribution of compounds in each sample. The two lines perpendicular to the X axis in the figure are 20%, 30% CV reference line, and the line parallel to the X axis is 60% of the reference line. Number of Compounds (CV<=30%): The number of compounds with a relative peak area CV of 30% or less in the QC sample. Ratio: The ratio of the number of compounds with a relative peak area CV less than or equal to 30% in the QC sample to the total number of compounds detected. Ratio >=60%, the data quality is qualified.

**Supplementary figure S3.** Alpha diversity at the phylum and genus level. *p < 0.05, **p < 0.01, ***p < 0.001

**Supplementary figure S4.** The Wilcoxon rank-sum test in HCC and non-HCC. (A) At the phylum level. (B) At the genus level. (C) At the phylum level. Only the relative abundance of top30 is given in the figure.

**Supplementary figure S5.** Serum metabolite changes in LC and NC. (A) PLS-DA shows the differences between LC and NC. The abscissa (PC1) and the ordinate (PC2) are the two main coordinates that explain the greatest difference between samples. The number is the score of the principal component, which represents the percentage of the explanation on overall variance of the specific pricipal component. The graph’s points represent samples, and different colors represent various sample grouping information; similar samples are clustered together. (B) The two rightmost points in the figure are the actual R2Y and Q2 values of the model, and the remaining points are the R2Y and Q2 values obtained by randomly arranging the samples used. This result is mainly used to judge whether the model is overfit and the validity of the model. (C) Volcano Plot differ between LC and NC.Green is the down-regulated differential metabolite (labeled green), red is the up-regulated differential metabolite (labeled red), and metabolites without difference are labeled purple-gray. (D) The KSM panel had AUC values of 0.93±0.12 in LC versus NC. KGM, key gut microbes; KSM, key serum metabolites; ROC, receiver operating characteristic; The abscissa of the ROC curve is the false positive rate, the ordinate is the true positive rate, the blue curve is the average curve after 10 folds, the AUC is the area under the curve, and the shaded region is the upper and lower 1 standard deviation.

**Supplementary figure S6.** (A) The heat map of top 20 differential species and differential metabolites with the smallest p-values for every omics in LC vs. NC. Columns represent the differential metabolites and rows represent the differential species. The color blocks represent the correlation coefficient. The darker the color, the stronger the correlation between the different species and the different metabolites. Red represents positive correlation and blue represents negative correlation. * represents p<0.05, ** represents p<0.01. (B) The ROC curves of Random Forest analysis of species and metabolism biomarkers. Random forest ROC map of species and metabolome (ROC map of metabolome is on the left, ROC map of species is in the middle, and ROC map of species and metabolome is on the right).

## References

1. Bray F, Ferlay J, Soerjomataram I, et al. Global cancer statistics 2018:GLOBOCAN estimates of incidence and mortality worldwide for 36 cancers in 185 countries. CA Cancer J Clin. 2018;68:394–424.

2. Torre LA, Bray F, Siegel RL, Ferlay J, Lortet-Tieulent J, Jemal A. Global cancer statistics, 2012. CA Cancer J Clin 2015;65:87–108.

3. European association for the study of the liver, Electronic address EEE. European association for the study of the L EASL clinical practice guidelines: management of hepatocellular carcinoma. J Hepatol 2018;69:182–236.

4. Kobayashi t, aikata H, Kobayashi t, et al. Patients with early recurrence of hepatocellular carcinoma have poor prognosis. Hepatobiliary Pancreat Dis Int 2017;16:279–88.

5. Marrero JA, Lok ASF. Newer markers for hepatocellular carcinoma. Gastroenterology 2004;127(5 Suppl. 1):S113–S119.

6. Dodd D, Spitzer MH, Van Treuren W, Merrill BD, Hryckowian AJ, Higginbottom SK, Le A, Cowan TM, Nolan GP, Fischbach MA, Sonnenburg JL. A gut bacterial pathway metabolizes aromatic amino acids into nine circulating metabolites. Nature. 2017 Nov 30;551(7682):648–652.

7. Qin J, Li Y, Cai Z, Li S, Zhu J, Zhang F, Liang S, Zhang W, Guan Y, Shen D, Peng Y, Zhang D, Jie Z, Wu W, Qin Y, Xue W, Li J, Han L, Lu D, Wu P, Dai Y, Sun X, Li Z, Tang A, Zhong S, Li X, Chen W, Xu R, Wang M, Feng Q, Gong M, Yu J, Zhang Y, Zhang M, Hansen T, Sanchez G, Raes J, Falony G, Okuda S, Almeida M, LeChatelier E, Renault P, Pons N, Batto JM, Zhang Z, Chen H, Yang R, Zheng W, Li S, Yang H, Wang J, Ehrlich SD, Nielsen R, Pedersen O, Kristiansen K, Wang J. A metagenome-wide association study of gut microbiota in type 2 diabetes. Nature. 2012 Oct 4;490(7418):55–60.

8. C. Lucas, N. Barnich, and H. T. T. Nguyen, “Microbiota, inflammation and colorectal cancer,” International Journal of Molecular Sciences, vol. 18, no. 6, p. 1310, 2017.

9. Tripathi A, Debelius J, Brenner DA, et al. The gut-liver axis and the intersection with the microbiome. Nat Rev Gastroenterol Hepatol. 2018;15: 397–411.

10. Ponziani FR, Bhoori S, Castelli C, et al. Hepatocellular carcinoma is associated with gut microbiota profile and inflammation in nonalcoholic fatty liver disease. Hepatology. 2019;69:107–20.

11. Shen F, Zheng rD, Sun XQ, et al. gut microbiota dysbiosis in patients with non-alcoholic fatty liver disease. Hepatobiliary Pancreat Dis Int 2017;16:375–81.

12. Bajaj JS, Heuman DM, Hylemon PB, et al. altered profile of human gut microbiome is associated with cirrhosis and its complications. J Hepatol 2014;60:940–7.

13. Qin J, li Y, cai Z, et al. a metagenome-wide association study of gut microbiota in type 2 diabetes. Nature 2012;490:55–60.

14. Yu J, Feng Q, Wong SH, et al. Metagenomic analysis of faecal microbiome as a tool towards targeted non-invasive biomarkers for colorectal cancer. Gut 2017;66:70–8.

15. ren Z, Jiang J, Xie H, et al. gut microbial profile analysis by MiSeq sequencing of pancreatic carcinoma patients in china. Oncotarget 2017;8.

16. Ren Z, Li A, Jiang J, Zhou L, Yu Z, Lu H, Xie H, Chen X, Shao L, Zhang R, Xu S, Zhang H, Cui G, Chen X, Sun R, Wen H, Lerut JP, Kan Q, Li L, Zheng S. Gut microbiome analysis as a tool towards targeted non-invasive biomarkers for early hepatocellular carcinoma. Gut. 2019 Jun;68(6):1014–1023.

17. Huang H, Ren Z, Gao X, Hu X, Zhou Y, Jiang J, Lu H, Yin S, Ji J, Zhou L, Zheng S. Integrated analysis of microbiome and host transcriptome reveals correlations between gut microbiota and clinical outcomes in HBV-related hepatocellular carcinoma. Genome Med. 2020 Nov 23;12(1):102.

18. Louis P, Hold GL, Flint HJ. The gut microbiota, bacterial metabolites and colorectal cancer. Nat Rev Microbiol. 2014; 12: 661–72.

19. Anand S, Kaur H, Mande SS. Comparative In silico Analysis of Butyrate Production Pathways in Gut Commensals and Pathogens. Front Microbiol. 2016; 7: 1945.

20. O’Keefe SJ. Diet, microorganisms and their metabolites, and colon cancer. Nat Rev Gastroenterol Hepatol. 2016; 13: 691–706.

21. Campbell, I. (2006). Liver: metabolic functions. Anaesth. Intensive Care Med.7, 51–54.

22. Beyoğlu D, Idle JR. The metabolomic window into hepatobiliary disease. J Hepatol 2013;59:842–58.

23. Al- Zoughbi W, Al- Zhoughbi W, Huang J, Paramasivan GS, et al. Tumor macroenvironment and metabolism. Semin Oncol 2014;41:281–95.

24. Vander Heiden MG, DeBerardinis RJ. Understanding the intersections between metabolism and cancer biology. Cell 2017;168:657–69.

25. Patel S, Ahmed S. Emerging field of metabolomics: big promise for cancer biomarker identification and drug discovery. J Pharm Biomed Anal 2015;107:63–74.

26. Tong M, Wong TL, Zhao H, Zheng Y, Xie YN, Li CH, Zhou L, Che N, Yun JP, Man K, Lee TK, Cai Z, Ma S. Loss of tyrosine catabolic enzyme HPD promotes glutamine anaplerosis through mTOR signaling in liver cancer. Cell Rep. 2021 Aug 24;36(8):109617.

27. Liu J, Geng W, Sun H, Liu C, Huang F, Cao J, Xia L, Zhao H, Zhai J, Li Q, Zhang X, Kuang M, Shen S, Xia Q, Wong VW, Yu J. Integrative metabolomic characterisation identifies altered portal vein serum metabolome contributing to human hepatocellular carcinoma. Gut. 2022 Jun;71(6):1203–1213.

28. SOAP: short oligonucleotide alignment program Ruiqiang Li, Yingrui Li, Karsten Kristiansen, Jun Wang Bioinformatics. 2008 Mar 1;24(5):713–4.

29. Fast gapped-read alignment with Bowtie 2 Ben Langmead, Steven L Salzberg Nat Methods. 2012 Mar 4;9(4):357–9.

30. MEGAHIT: an ultra-fast single-node solutionfor large and complex metagenomics assembly via succinct de Bruijn graph Dinghua Li, Chi-Man Liu, Ruibang Luo, Kunihiko Sadakane, Tak-Wah Lam Bioinformatics. 2015 May 15;31(10):1674–6.

31. Ab initio gene identification in metagenomic sequences Wenhan Zhu, Alexandre Lomsadze, Mark Borodovsky Nucleic Acids Res. 2010 Jul;38(12):e132. doi: 10.1093/nar/gkq275. Epub 2010 Apr 19.

32. CD-HIT: accelerated for clustering the next-generation sequencing data Limin Fu, Beifang Niu, Zhengwei Zhu, Sitao Wu, Weizhong Li Bioinformatics. 2012 Dec 1;28(23):3150–2.

33. Salmon provides fast and bias-aware quantification of transcript expression Rob Patro, Geet Duggal, Michael I Love, Rafael A Irizarry, Carl Kingsford Nat Methods. 2017 Apr;14(4):417–419.

34. Fast and sensitive protein alignment using DIAMOND Benjamin Buchfink, Chao Xie, Daniel H Huson Nature Methods. 2015 Jan;12(1):59–60.

35. Dunn W B, Broadhurst D, Begley P, et al. Procedures for large-scale metabolic profiling of serum and plasma using gas chromatography and liquid chromatography coupled to mass spectrometry[J]. Nature Protocols, 2011, 6(7):1060–1083.

36. Sarafian M H, Gaudin M, Lewis M R, et al. Objective Set of Criteria for Optimization of Sample Preparation Proced ures for Ultra-High Throughput Untargeted Blood Plasma Lipid Profiling by Ultra Performance Liquid Chromatography–Mass Spectrometry[J]. Analytical Chemistry, 2014, 86(12):5766–5774.

37. The distribution of the thermally tolerant symbiont lineage (Symbiodinium clade D) in corals from Hawaii: correlations with host and the history of ocean thermal stress Michael Stat, Xavier Pochon, Erik C Franklin, John F Bruno, Kenneth S Casey, Elizabeth R Selig, Ruth D Gates Ecol Evol. 2013 May;3(5):1317–29.

38. Non-parametric multivariate analyses of changes in community structure K. Robert Clarke Austral Ecology. 1993 Mar;18 (1): 117–143.

39. Wen, B., et al.metaX: a flexible and comprehensive software for processing metabolomics data. BMC Bioinformatics 2017,18(1): 183.

40. Ren Z, Li A, Jiang J, Zhou L, Yu Z, Lu H, Xie H, Chen X, Shao L, Zhang R, Xu S, Zhang H, Cui G, Chen X, Sun R, Wen H, Lerut JP, Kan Q, Li L, Zheng S. Gut microbiome analysis as a tool towards targeted non-invasive biomarkers for early hepatocellular carcinoma. Gut. 2019 Jun;68(6):1014–1023. doi: 10.1136/gutjnl-2017-315084. Epub 2018 Jul 25. PMID: 30045880; PMCID: PMC6580753.

41. Wexler HM. Bacteroides: the good, the bad, and the nitty-gritty. Clin Microbiol Rev. 2007 Oct;20(4):593–621. doi: 10.1128/CMR.00008-07. PMID: 17934076; PMCID: PMC2176045.

42. Wang H, Wang Q, Yang C, Guo M, Cui X, Jing Z, Liu Y, Qiao W, Qi H, Zhang H, Zhang X, Zhao N, Zhang M, Chen M, Zhang S, Xu H, Zhao L, Qiao M, Wu Z. Bacteroides acidifaciens in the gut plays a protective role against CD95-mediated liver injury. Gut Microbes. 2022 Jan-Dec;14(1):2027853. doi: 10.1080/19490976.2022.2027853. PMID: 35129072; PMCID: PMC8820816.

43. Huang H, Ren Z, Gao X, Hu X, Zhou Y, Jiang J, Lu H, Yin S, Ji J, Zhou L, Zheng S. Integrated analysis of microbiome and host transcriptome reveals correlations between gut microbiota and clinical outcomes in HBV-related hepatocellular carcinoma. Genome Med. 2020 Nov 23;12(1):102. doi: 10.1186/s13073-020-00796-5. PMID: 33225985; PMCID: PMC7682083.

44. Zhen H, Qian X, Fu X, Chen Z, Zhang A, Shi L. Regulation of Shaoyao Ruangan Mixture on Intestinal Flora in Mice With Primary Liver Cancer. Integr Cancer Ther. 2019 Jan-Dec;18:1534735419843178. doi: 10.1177/1534735419843178. PMID: 31006277; PMCID: PMC6477757.

45. Ram AK, Vairappan B, Srinivas BH. Nimbolide attenuates gut dysbiosis and prevents bacterial translocation by improving intestinal barrier integrity and ameliorating inflammation in hepatocellular carcinoma. Phytother Res. 2022 May;36(5):2143–2160. doi: 10.1002/ptr.7434. Epub 2022 Mar 1. PMID: 35229912.

46. Kim SS, Eun JW, Cho HJ, Song DS, Kim CW, Kim YS, Lee SW, Kim YK, Yang J, Choi J, Yim HJ, Cheong JY. Microbiome as a potential diagnostic and predictive biomarker in severe alcoholic hepatitis. Aliment Pharmacol Ther. 2021 Feb;53(4):540–551. doi: 10.1111/apt.16200. Epub 2020 Dec 2. PMID: 33264437.

47. Wei Y, Li Y, Yan L, Sun C, Miao Q, Wang Q, Xiao X, Lian M, Li B, Chen Y, Zhang J, Li Y, Huang B, Li Y, Cao Q, Fan Z, Chen X, Fang JY, Gershwin ME, Tang R, Ma X. Alterations of gut microbiome in autoimmune hepatitis. Gut. 2020 Mar;69(3):569–577. doi: 10.1136/gutjnl-2018-317836. Epub 2019 Jun 14. PMID: 31201284.

48. Zhang L, Wu YN, Chen T, Ren CH, Li X, Liu GX. Relationship between intestinal microbial dysbiosis and primary liver cancer. Hepatobiliary Pancreat Dis Int. 2019 Apr;18(2):149–157. doi: 10.1016/j.hbpd.2019.01.002. Epub 2019 Jan 4. PMID: 30661942.

49. Tang Y, Zhou H, Xiang Y, Cui F. The diagnostic potential of gut microbiome for early hepatitis B virus-related hepatocellular carcinoma. Eur J Gastroenterol Hepatol. 2021 Dec 1;33(1S Suppl 1):e167–e175. doi: 10.1097/MEG.0000000000001978. PMID: 33208683.

50. Mao J, Wang D, Long J, Yang X, Lin J, Song Y, Xie F, Xun Z, Wang Y, Wang Y, Li Y, Sun H, Xue J, Song Y, Zuo B, Zhang J, Bian J, Zhang T, Yang X, Zhang L, Sang X, Zhao H. Gut microbiome is associated with the clinical response to anti-PD-1 based immunotherapy in hepatobiliary cancers. J Immunother Cancer. 2021 Dec;9(12):e003334. doi: 10.1136/jitc-2021-003334. PMID: 34873013; PMCID: PMC8650503.

51. Okubo R, Kinoshita T, Katsumata N, Uezono Y, Xiao J, Matsuoka YJ. Impact of chemotherapy on the association between fear of cancer recurrence and the gut microbiota in breast cancer survivors. Brain Behav Immun. 2020 Mar;85:186–191. doi: 10.1016/j.bbi.2019.02.025. Epub 2019 Feb 25. PMID: 30818031.

52. Ishaq HM, Mohammad IS, Hussain R, Parveen R, Shirazi JH, Fan Y, Shahzad M, Hayat K, Li H, Ihsan A, Muhammad KS, Usman M, Zhang S, Yuan L, Ullah S, Paiva-Santos AC, Xu J. Gut-Thyroid axis: How gut microbial dysbiosis associated with euthyroid thyroid cancer. J Cancer. 2022 Mar 28;13(6):2014–2028. doi: 10.7150/jca.66816. PMID: 35399732; PMCID: PMC8990431.

53. Zhao SS, Chen L, Yang J, Wu ZH, Wang XY, Zhang Q, Liu WJ, Liu HX. Altered Gut Microbial Profile Accompanied by Abnormal Fatty Acid Metabolism Activity Exacerbates Endometrial Cancer Progression. Microbiol Spectr. 2022 Oct 13:e0261222. doi: 10.1128/spectrum.02612-22. Epub ahead of print. PMID: 36227107.

54. Chen Y, Ma J, Dong Y, Yang Z, Zhao N, Liu Q, Zhai W, Zheng J. Characteristics of Gut Microbiota in Patients With Clear Cell Renal Cell Carcinoma. Front Microbiol. 2022 Jul 4;13:913718. doi: 10.3389/fmicb.2022.913718. PMID: 35865926; PMCID: PMC9295744.

55. Lima SF, Gogokhia L, Viladomiu M, Chou L, Putzel G, Jin WB, Pires S, Guo CJ, Gerardin Y, Crawford CV, Jacob V, Scherl E, Brown SE, Hambor J, Longman RS. Transferable Immunoglobulin A-Coated Odoribacter splanchnicus in Responders to Fecal Microbiota Transplantation for Ulcerative Colitis Limits Colonic Inflammation. Gastroenterology. 2022 Jan;162(1):166–178. doi: 10.1053/j.gastro.2021.09.061. Epub 2021 Oct 2. PMID: 34606847; PMCID: PMC8678328.

56. Wang J, Qie J, Zhu D, Zhang X, Zhang Q, Xu Y, Wang Y, Mi K, Pei Y, Liu Y, Ji G, Liu X. The landscape in the gut microbiome of long-lived families reveals new insights on longevity and aging - relevant neural and immune function. Gut Microbes. 2022 Jan-Dec;14(1):2107288. doi: 10.1080/19490976.2022.2107288. PMID: 35939616; PMCID: PMC9361766.

57. Xing C, Wang M, Ajibade AA, Tan P, Fu C, Chen L, Zhu M, Hao ZZ, Chu J, Yu X, Yin B, Zhu J, Shen WJ, Duan T, Wang HY, Wang RF. Microbiota regulate innate immune signaling and protective immunity against cancer. Cell Host Microbe. 2021 Jun 9;29(6):959–974.e7. doi: 10.1016/j.chom.2021.03.016. Epub 2021 Apr 23. PMID: 33894128; PMCID: PMC8192480.

58. Yoshimoto S, Loo TM, Atarashi K, Kanda H, Sato S, Oyadomari S, Iwakura Y, Oshima K, Morita H, Hattori M, Honda K, Ishikawa Y, Hara E, Ohtani N. Obesity-induced gut microbial metabolite promotes liver cancer through senescence secretome. Nature. 2013 Jul 4;499(7456):97–101. doi: 10.1038/nature12347. Epub 2013 Jun 26. Erratum in: Nature. 2014 Feb 20;506(7488):396. Hattori, Masahisa [corrected to Hattori, Masahira]. PMID: 23803760.

59. Quante M, Bhagat G, Abrams JA, Marache F, Good P, Lee MD, Lee Y, Friedman R, Asfaha S, Dubeykovskaya Z, Mahmood U, Figueiredo JL, Kitajewski J, Shawber C, Lightdale CJ, Rustgi AK, Wang TC. Bile acid and inflammation activate gastric cardia stem cells in a mouse model of Barrett-like metaplasia. Cancer Cell. 2012 Jan 17;21(1):36–51. doi: 10.1016/j.ccr.2011.12.004. PMID: 22264787; PMCID: PMC3266546.

60. Jia W, Xie G, Jia W. Bile acid-microbiota crosstalk in gastrointestinal inflammation and carcinogenesis. Nat Rev Gastroenterol Hepatol. 2018 Feb;15(2):111–128. doi: 10.1038/nrgastro.2017.119. Epub 2017 Oct 11. PMID: 29018272; PMCID: PMC5899973.

61. Lechner S, Müller-Ladner U, Schlottmann K, Jung B, McClelland M, Rüschoff J, Welsh J, Schölmerich J, Kullmann F. Bile acids mimic oxidative stress induced upregulation of thioredoxin reductase in colon cancer cell lines. Carcinogenesis. 2002 Aug;23(8):1281–8. doi: 10.1093/carcin/23.8.1281. PMID: 12151345.

62. Xie G, Wang X, Huang F, Zhao A, Chen W, Yan J, Zhang Y, Lei S, Ge K, Zheng X, Liu J, Su M, Liu P, Jia W. Dysregulated hepatic bile acids collaboratively promote liver carcinogenesis. Int J Cancer. 2016 Oct 15;139(8):1764–75. doi: 10.1002/ijc.30219. Epub 2016 Jun 17. PMID: 27273788; PMCID: PMC5493524.

63. Kühn T, Stepien M, López-Nogueroles M, Damms-Machado A, Sookthai D, Johnson T, Roca M, Hüsing A, Maldonado SG, Cross AJ, Murphy N, Freisling H, Rinaldi S, Scalbert A, Fedirko V, Severi G, Boutron-Ruault MC, Mancini FR, Sowah SA, Boeing H, Jakszyn P, Sánchez MJ, Merino S, Colorado-Yohar S, Barricarte A, Khaw KT, Schmidt JA, Perez-Cornago A, Trichopoulou A, Karakatsani A, Thriskos P, Palli D, Agnoli C, Tumino R, Sacerdote C, Panico S, Bueno-de-Mesquita B, van Gils CH, Heath AK, Gunter MJ, Riboli E, Lahoz A, Jenab M, Kaaks R. Prediagnostic Plasma Bile Acid Levels and Colon Cancer Risk: A Prospective Study. J Natl Cancer Inst. 2020 May 1;112(5):516–524. doi: 10.1093/jnci/djz166. Erratum in: J Natl Cancer Inst. 2020 Oct 1;112(10):1077. PMID: 31435679; PMCID: PMC7225675.

64. Farhat Z, Freedman ND, Sampson JN, Falk RT, Koshiol J, Weinstein SJ, Albanes D, Sinha R, Loftfield E. A prospective investigation of serum bile acids with risk of liver cancer, fatal liver disease, and biliary tract cancer. Hepatol Commun. 2022 Sep;6(9):2391–2399. doi: 10.1002/hep4.2003. Epub 2022 Jun 8. PMID: 35678016; PMCID: PMC9426378.

65. Liao M, Zhao J, Wang T, Duan J, Zhang Y, Deng X. Role of bile salt in regulating Mcl-1 phosphorylation and chemoresistance in hepatocellular carcinoma cells. Mol Cancer. 2011 Apr 20;10:44. doi: 10.1186/1476-4598-10-44. PMID: 21507240; PMCID: PMC3107804.

66. Gao L, Lv G, Li R, Liu WT, Zong C, Ye F, Li XY, Yang X, Jiang JH, Hou XJ, Jing YY, Han ZP, Wei LX. Glycochenodeoxycholate promotes hepatocellular carcinoma invasion and migration by AMPK/mTOR dependent autophagy activation. Cancer Lett. 2019 Jul 10;454:215–223. doi: 10.1016/j.canlet.2019.04.009. Epub 2019 Apr 11. PMID: 30980867.

67. Mackintosh C, Yuan C, Ou FS, Zhang S, Niedzwiecki D, Chang IW, O’Neil BH, Mullen BC, Lenz HJ, Blanke CD, Venook AP, Mayer RJ, Fuchs CS, Innocenti F, Nixon AB, Goldberg RM, O’Reilly EM, Meyerhardt JA, Ng K. Association of Coffee Intake With Survival in Patients With Advanced or Metastatic Colorectal Cancer. JAMA Oncol. 2020 Nov 1;6(11):1713–1721. doi: 10.1001/jamaoncol.2020.3938. PMID: 32940631; PMCID: PMC7499248.

68. Oh JK, Sandin S, Ström P, Löf M, Adami HO, Weiderpass E. Prospective study of breast cancer in relation to coffee, tea and caffeine in Sweden. Int J Cancer. 2015 Oct 15;137(8):1979–89. doi: 10.1002/ijc.29569. Epub 2015 Apr 30. PMID: 25885188.

69. Wilson KM, Kasperzyk JL, Rider JR, Kenfield S, van Dam RM, Stampfer MJ, Giovannucci E, Mucci LA. Coffee consumption and prostate cancer risk and progression in the Health Professionals Follow-up Study. J Natl Cancer Inst. 2011 Jun 8;103(11):876–84. doi: 10.1093/jnci/djr151. Epub 2011 May 17. PMID: 21586702; PMCID: PMC3110172.

70. Inoue M, Tsugane S. Coffee Drinking and Reduced Risk of Liver Cancer: Update on Epidemiological Findings and Potential Mechanisms. Curr Nutr Rep. 2019 Sep;8(3):182–186. doi: 10.1007/s13668-019-0274-1. PMID: 30977056.

71. Guertin KA, Loftfield E, Boca SM, Sampson JN, Moore SC, Xiao Q, Huang WY, Xiong X, Freedman ND, Cross AJ, Sinha R. Serum biomarkers of habitual coffee consumption may provide insight into the mechanism underlying the association between coffee consumption and colorectal cancer. Am J Clin Nutr. 2015 May;101(5):1000–11. doi: 10.3945/ajcn.114.096099. Epub 2015 Mar 11. PMID: 25762808; PMCID: PMC4409687.

72. Benoehr P, Krueth P, Bokemeyer C, Grenz A, Osswald H, Hartmann JT. Nephroprotection by theophylline in patients with cisplatin chemotherapy: a randomized, single-blinded, placebo-controlled trial. J Am Soc Nephrol. 2005 Feb;16(2):452–8. doi: 10.1681/ASN.2004030225. Epub 2004 Dec 8. PMID: 15590762.

73. Ye J, Mao L, Xie L, Zhang R, Liu Y, Peng L, Yang J, Li Q, Yuan M. Discovery of a Series of Theophylline Derivatives Containing 1,2,3-Triazole for Treatment of Non-Small Cell Lung Cancer. Front Pharmacol. 2021 Oct 26;12:753676. doi: 10.3389/fphar.2021.753676. PMID: 34764872; PMCID: PMC8576520.

74. Luo P, Yin P, Hua R, Tan Y, Li Z, Qiu G, Yin Z, Xie X, Wang X, Chen W, Zhou L, Wang X, Li Y, Chen H, Gao L, Lu X, Wu T, Wang H, Niu J, Xu G. A Large-scale, multicenter serum metabolite biomarker identification study for the early detection of hepatocellular carcinoma. Hepatology. 2018 Feb;67(2):662–675. doi: 10.1002/hep.29561. Epub 2018 Jan 2. PMID: 28960374; PMCID: PMC6680350.

