## Supplementary figures and images for "Integrated microbiome and metabolome analysis reveals the interaction between intestinal flora and serum metabolites as potential biomarkers in hepatocellular carcinoma patients"

### S1.jpg

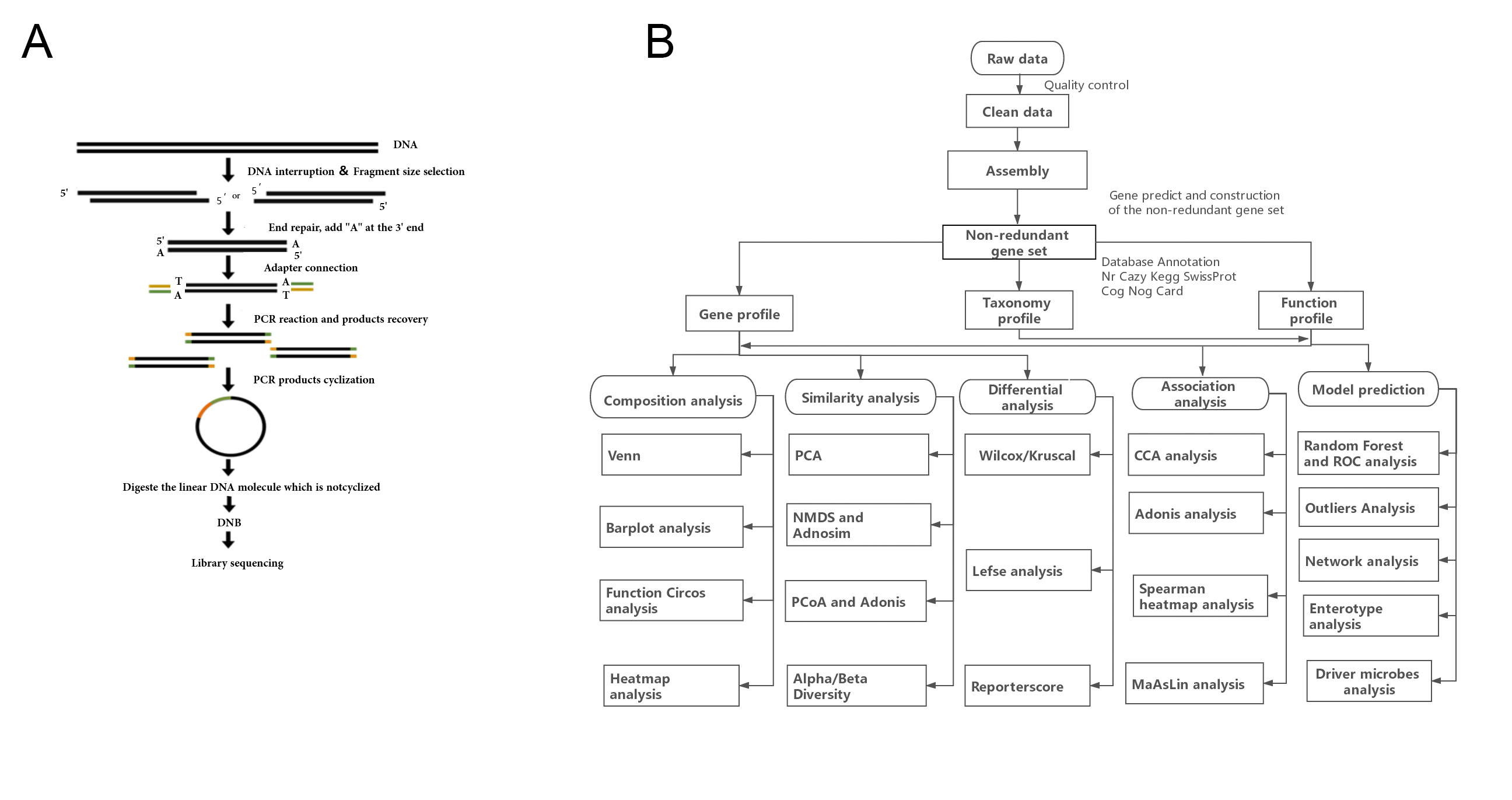

### S2.jpg

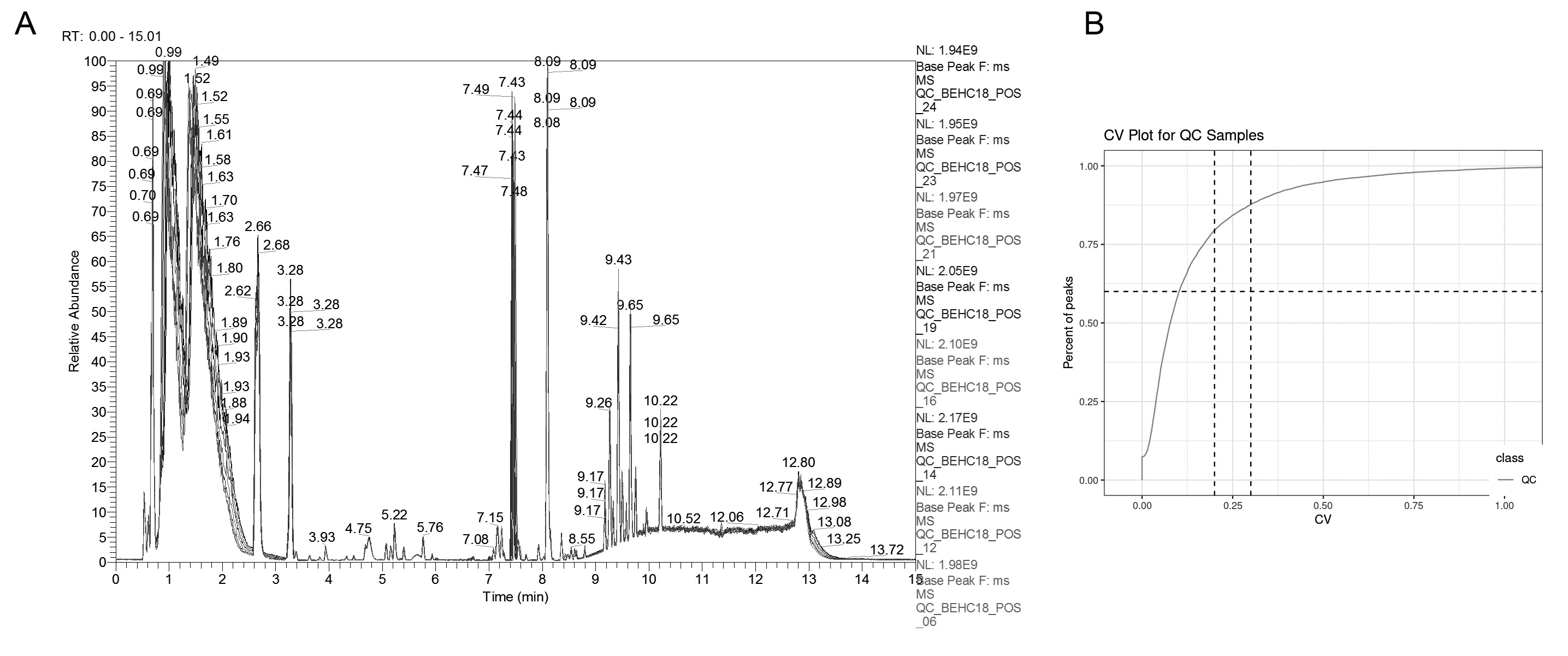

### s3.jpg

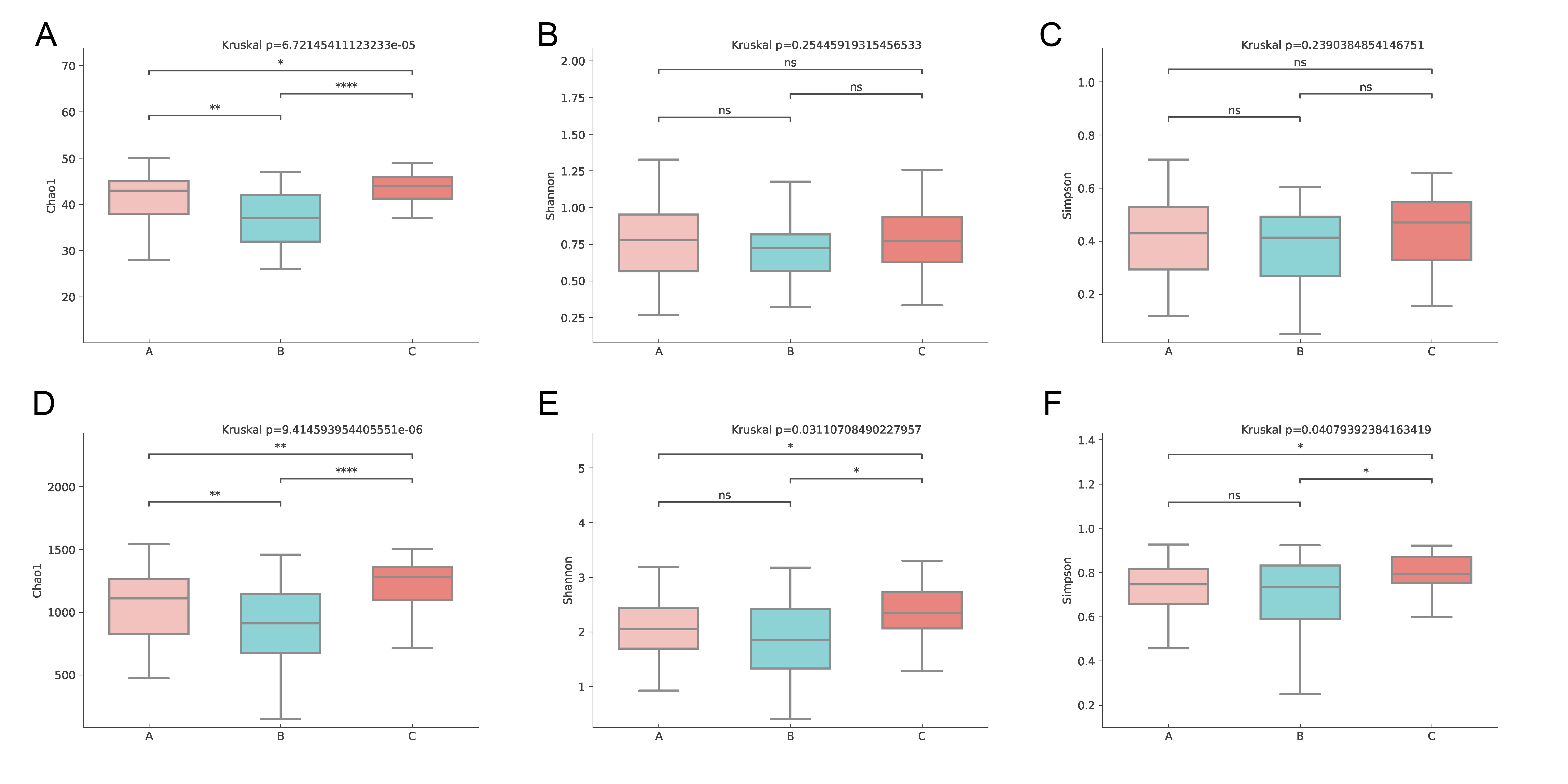

### S4.jpg

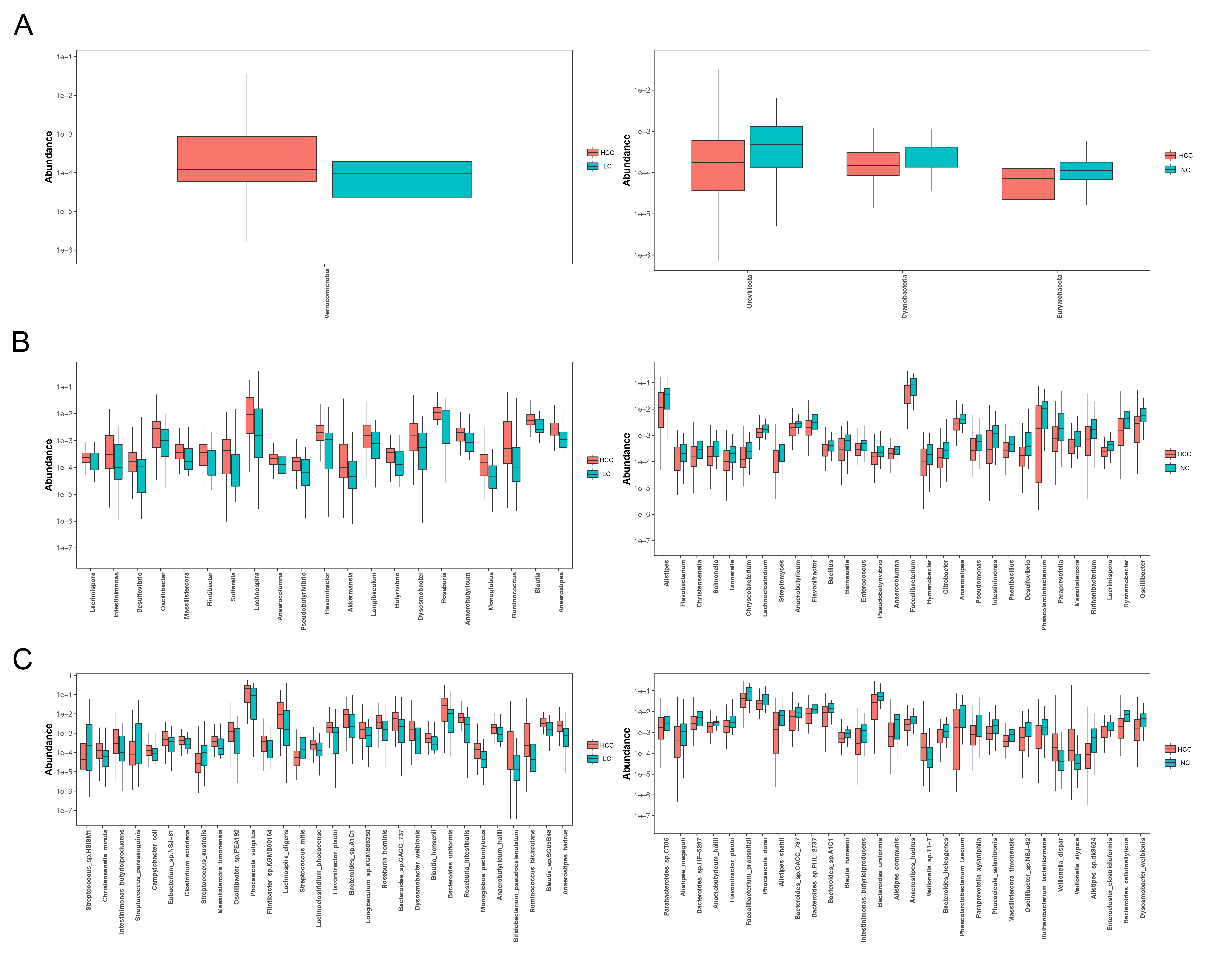

### S5.jpg

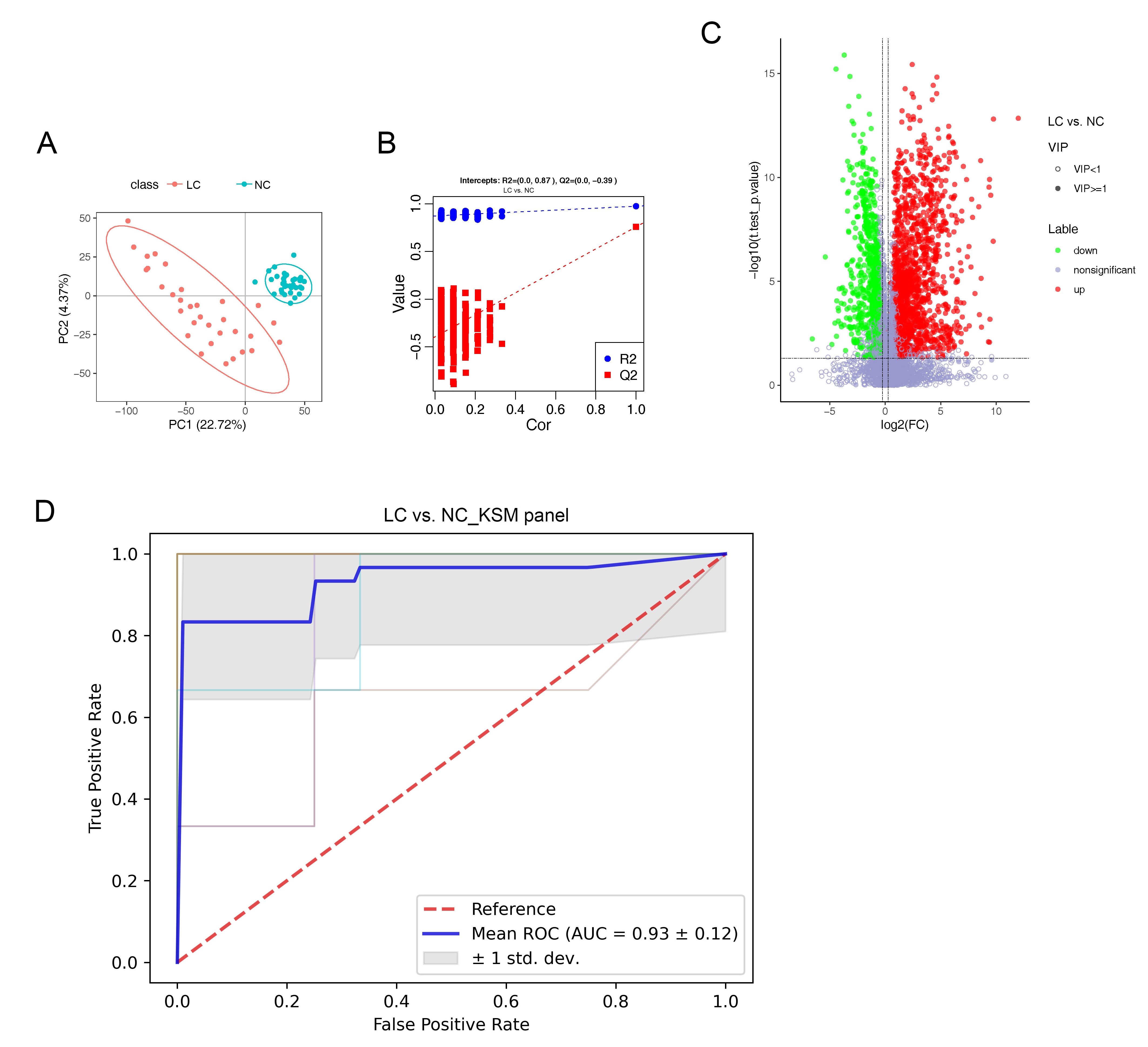

### S6.jpg

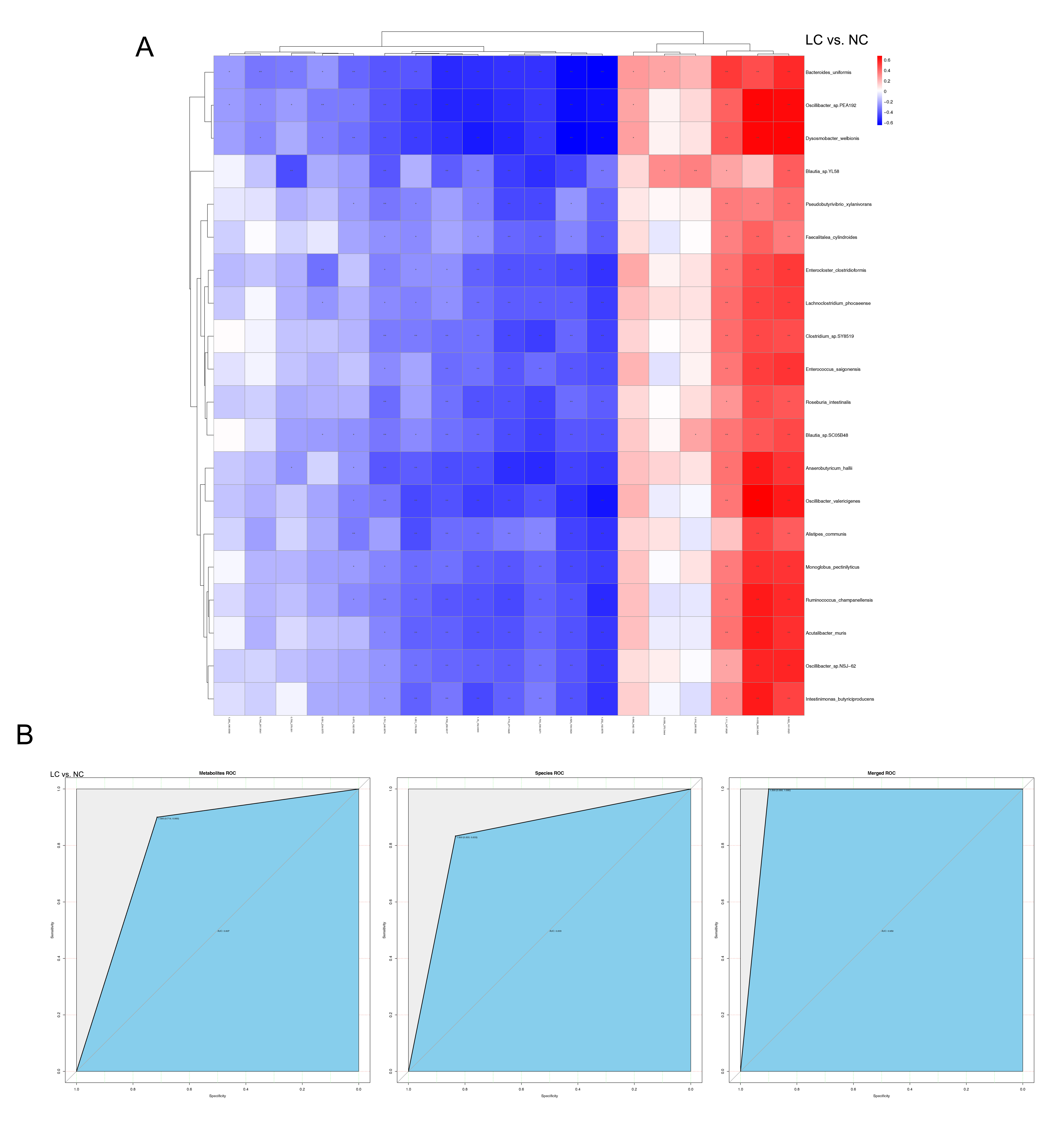
